# Cannabis and Tobacco Co-Use Predicts Psychosis in Clinical High Risk Cohorts

**DOI:** 10.1101/2025.09.19.25336202

**Authors:** Daniel Bello, Sophia H. Blyth, Rachel A. Rabin, Jean Addington, Carrie E. Bearden, Kristin Cadenhead, Tyrone D. Cannon, Ricardo E Carrión, Barbara Cornblatt, Matcheri Keshavan, Daniel H. Mathalon, Diana O. Perkins, Larry Seidman, William S. Stone, Ming T. Tsuang, Elaine F. Walker, Scott Woods, Roscoe O. Brady, Heather Burrell Ward

## Abstract

Cannabis and tobacco use are highly prevalent among people with psychosis and are associated with medical comorbidities and poor prognosis. Concurrent use of cannabis and tobacco (“co-use”) is rising in the general population but has not been studied in psychosis. Given the devastating consequences of cannabis and tobacco use, it is critical to understand how their co-use affects psychiatric symptoms and the development of psychosis. We used the North American Prodrome Longitudinal Study 2, a multi-site prospective study of individuals at clinical high risk for psychosis (CHR) and healthy controls, to examine baseline differences in psychiatric symptoms and conversion to psychosis across substance groups: 1) CHR tobacco use, 2) CHR cannabis use, 3) CHR co-use, 4) CHR non-tobacco or cannabis substance use, 5) CHR without substance use, and 6) healthy controls. Among 1,014 participants (734 CHR, 280 controls), more frequent cannabis and tobacco use was linked to greater psychiatric symptom severity, including psychosis, anxiety, and depression. In survival analyses, co-use (HR *=* 2.53, 95% CI [1.44–4.45], *p* =.001), especially heavy co-use (HR = 3.63, 95% CI: 1.53–8.63, *p* = 0.003), was associated with increased risk of conversion to psychosis. Co-use of tobacco and cannabis was not associated with psychiatric symptom severity but did predict higher risk of conversion to psychosis. The combination of cannabis and tobacco use may exert a synergistic effect, amplifying conversion risk more than either substance alone, or may be a marker of an elevated underlying psychosis risk. These results highlight the need for early intervention strategies that address co-use in CHR populations to mitigate potential long-term psychiatric consequences.

## Introduction

Cannabis and tobacco use are highly prevalent among people with psychosis.^1^ Up to 60% of people with schizophrenia use tobacco – a prevalence three times that of the general population. Cannabis use is also common, especially among individuals with first episode psychosis.^2^ The current and lifetime rates of cannabis use disorders among people with psychosis are estimated to be 16% and 27.1%, respectively.^3^ In schizophrenia, 43% have a cannabis use disorder, compared to 15% in the population.^4^

Tobacco and cannabis use are associated with significant medical and psychiatric comorbidities in people with psychosis. Tobacco use is the leading preventable cause of early mortality in schizophrenia, leading to a 20-year decreased life expectancy compared to the general population.^5,6^ Individuals with psychosis smoke more heavily, are more nicotine dependent, and are less likely to quit smoking than people without a psychotic disorder.^6–8^

Cannabis use is associated with psychotic symptom exacerbation, psychotic relapse, treatment nonadherence, and poorer overall functioning.^4,9–11^ Lifetime cannabis use is associated with a 1.4-fold increase in the risk of developing psychotic illness, with cannabis dependence conferring a 3.4-fold increase.^10^ Cannabis use is involved in roughly half of psychotic disorder cases, and regular use is a predictor of heightened schizophrenia risk.^12–15^ Cannabis use is known to cause positive symptoms of psychosis, including paranoia, hallucinations,^16^ as well as anhedonia and amotivation,^17,18^ which are commonly associated with negative symptoms.

Substance use generally begins in adolescence, prior to the onset of psychosis. Approximately 77% of individuals report using tobacco before the onset of psychotic symptoms.^6^ Similarly, individuals who use cannabis have onset of schizophrenia 2-3 years earlier than those who do not use cannabis, which is predictive of poorer long-term clinical outcomes.^10^

Existing literature has studied the effects of single substances (i.e., tobacco or cannabis use) on psychiatric symptoms and treatment outcomes in psychosis. However, the real world is not a controlled trial of tobacco or cannabis use alone, as many individuals use both substances.

Concurrent use of tobacco and cannabis, defined as “co-use,” has become increasingly prevalent in the general population. Co-use can include using the substances at the same time, on the same occasion, or within a defined timeframe where their effects may overlap. Co-use of tobacco and cannabis can occur *simultaneously*, such as spliffs, which are joints that include cannabis and loose-leaf tobacco, or *asynchronously*, where the individual uses both substances at different times (e.g., using tobacco in the morning and cannabis at night).^12^ Of individuals aged 18-25 in the United States, 20% of individuals who smoke cigarettes also use cannabis daily. In 2014, the prevalence of daily cannabis use among cigarette smokers doubled from 4.9% in 2002 to 9%.^12^ Importantly, co-use of tobacco and cannabis is associated with higher rates of psychiatric conditions, worse substance use outcomes, and poorer physical health.^12^ Both cannabis and tobacco may sensitize dopamine neurotransmission, cortical maturation, and genetic risk,^13^ thereby amplifying psychosis risk.^14,19^ Nicotine use may predispose to cannabis use or potentiate its effects by increasing tetrahydrocannabinol (THC) absorption.^20^

Given the devastating medical and psychiatric consequences of use of these substances, it is critical to understand the effects of co-use on psychiatric symptoms and risk of developing a psychotic disorder. Because substance use often begins prior to the onset of psychosis, we investigated patterns of tobacco and cannabis co-use during the clinical high risk (CHR) period, a time prior to the onset of psychosis when individuals experience attenuated psychotic symptoms, alterations in mood and anxiety, and changes in social functioning.^21,22^ We compared baseline symptom severity among CHR with current co-use, tobacco use only or cannabis use only, other substance use, and no substance use from the North American Prodrome Longitudinal Study (NAPLS2), a multisite prospective study of individuals at risk for psychosis who underwent neuroimaging and clinical characterization then were followed for 2 years for development of a psychotic disorder (i.e., conversion to psychosis). We also assessed if baseline tobacco and cannabis co-use was associated with conversion to psychosis. We hypothesized that CHR with co-use would have 1) more severe psychiatric symptoms and 2) a higher rate of conversion to psychosis than CHR using only tobacco or cannabis.

## Materials & Methods

### Participants

NAPLS2 is a longitudinal case-control study studying individuals at CHR for psychosis across 8 sites in North America. Individuals meeting CHR criteria (N=734) and healthy controls (N=280) were enrolled. See supplement for inclusion/exclusion criteria. Participants underwent assessment at baseline and every 6 months for two years and upon conversion to psychosis (if applicable) from January 2009 to April 2013. Only baseline assessments were used for this analysis. Prior to participation, all participants provided written informed consent (or, if under age 18, informed assent with parental consent) in accordance with institutional review boards. See supplement for details.

### Measures

*Diagnosis*: CHR individuals met Criteria for the Psychosis Risk Syndrome^23^ based on the Structured Interview for Psychosis-Risk Syndrome (SIPS).^23^ Conversion to psychosis was determined from follow-up SIPS interview, defined as meeting the Presence of Psychosis Syndrome criteria.

*Substance Use*: Current substance use at baseline was measured using the Alcohol Use Scale/Drug Use Scale (AUS/DUS), which assesses substance use frequency over the past 30 days on an ordinal scale.^24^ Tobacco use was rated in cigarettes per day (0 = no use, 1 = occasionally, 2 = <10 per day, 3 = 11-25 per day, 4 = >25 per day). Cannabis frequency was reported as (0 = no use, 1 = once or twice per month, 2 = 3-4 times per month, 3 = 1-2 times per week, 4 = 3-4 times per week, 5 = almost daily). See supplement for details. We categorized CHR individuals into groups based on their reported substance use in the past 30 days: 1) Tobacco only; 2) Cannabis only; 3) Tobacco and Cannabis Co-Use; 4) Neither Tobacco nor Cannabis (Non-TC use); or 5) No substance use. Healthy controls had minimal substance use and so were treated as one group. Because these co-use categories did not consider intensity of use, tobacco and cannabis use were also treated as ordinal variables to test for dose-response relationships.

*Psychiatric Symptoms*: Severity of psychosis symptoms at baseline was rated using the Scale of Psychosis-Risk Symptoms (SOPS).^23^ Anxiety and depressive symptoms were measured with the Self-Rating Anxiety Scale (SAS),^25^ Social Interaction Anxiety Scale (SIAS),^26^ and Calgary Depression Scale for Schizophrenia (CDSS).^27^

### Statistical Approach

T-tests were used to compare continuous outcomes based on dichotomous variables. ANOVAs were used to compare continuous outcomes (e.g., symptom scores) based on three or more groups. Kruskal-Wallis tests were used to compare differences in ordinal variables (e.g., substance use frequency) across substance use groups. Spearman correlations were used to determine relationships with ordinal variables. Linear regression models were used to predict 1) symptom severity and 2) substance use frequency based on age, sex, site, diagnosis, and diagnosis*substance use interactions. All analyses were conducted in RStudio (Version 2023.03.1+446) using alpha<.05, except as indicated in results to correct for multiple comparisons.

### Survival Analyses

Cox proportional hazards regression models were used to examine the association between tobacco and cannabis use and conversion to psychosis. Time was measured as days from initial assessment to last follow up assessment or post-conversion assessment. We fitted five Cox proportional hazards models controlling for age and sex:

1. Cannabis use (ordinal)
2. Tobacco use (ordinal)
3. Categorical Co-Use (No use, Tobacco use only, Cannabis use only, Co-Use)
4. Categorical Intensity of Co-Use (No use, Light use, Heavy use for Tobacco and Cannabis)
5. Cannabis and Tobacco simultaneously (both ordinal)

The reference category for the categorical models was no use of either substance. Hazard ratios with 95% confidence intervals were calculated for each predictor. All analyses were conducted using R version 4.5.1 (2025-06-13) and the following packages: *survival* and *survminer*.

## Results

There were 1,014 participants with data for analysis (Supplemental Table 1). In the CHR group (n=734), 83 converted to psychosis (i.e. developed a psychotic disorder, CHR-C) during the 2-year study duration. Healthy Controls (n=280) were older than CHR individuals (19.73 vs. 18.50 years, t(1042) =-4.04, p < 0.001). There were no other differences in age, sex, or race. There were no differences in age, sex, race, or socioeconomic status between the CHR substance use groups (Table 1, Supplemental Figure 1).

**Table 1.** Demographics by Substance Use Group.

|  | Clinical High Risk (n=734) |  |  |  |  | Healthy Control (n=280) |
| --- | --- | --- | --- | --- | --- | --- |
|  | Co-Use (n=97) | Tobacco Use (n=78) | Cannabis Use (n=80) | No Tobacco or Cannabis Use (n=105) | No Substance Use (n=404) |  |
| **Age, y (SD)<sup>+</sup>** | 19.3 (3.0) | 20.3 (4.3) | 19.6 (3.8) | 21.2 (4.7) | 17.0 (3.8) | 19.7 (4.7) |
| **Sex, Male (%)<sup>++</sup>** | 69 (71.1) | 49 (62.8) | 46 (57.5) | 52 (49.5) | 220 (54.5) | 141 (50.4) |
| **Race (%)<sup>+++</sup>** |  |  |  |  |  |  |
| First Nations | 2 (2.1) | 0 (0.0) | 1 (1.3) | 0 (0.0) | 10 (2.7) | 4 (1.4) |
| East Asian | 1 (1.0) | 1 (1.3) | 4 (5.1) | 1 (1.0) | 12 (3.2) | 15 (5.4) |
| Southeast Asian | 0 (0.0) | 2 (2.6) | 0 (0.0) | 0 (0.0) | 13 (3.5) | 7 (2.5) |
| South Asian | 1 (1.0) | 2 (2.6) | 1 (1.3) | 2 (2.0) | 13 (3.5) | 8 (2.9) |
| Black | 13 (13.4) | 9 (11.5) | 12 (15.2) | 17 (16.2) | 58 (15.5) | 49 (17.5) |
| Central/South American | 1 (1.0) | 4 (5.1) | 2 (2.5) | 4 (3.8) | 21 (5.6) | 13 (4.6) |
| West/Central Asia and Middle East | 1 (1.0) | 2 (2.6) | 0 (0.0) | 0 (0.0) | 3 (0.8) | 2 (0.7) |
| White | 70 (72.2) | 49 (62.8) | 52 (65.8) | 64 (61.0) | 191 (51.1) | 152 (54.3) |
| Native Hawaiian or Pacific Islander | 0 (0.0) | 0 (0.0) | 0 (0.0) | 0 (0.0) | 2 (0.5) | 1 (0.4) |
| Interracial | 8 (8.3) | 9 (11.5) | 7 (8.9) | 17 (16.2) | 51 (13.6) | 29 (10.4) |
| **Hispanic (%)<sup>++++</sup>** | 9 (9.3) | 8 (10.3) | 12 (15.0) | 22 (21.0) | 91 (22.6) | 50 (17.9) |
| **Taking Antipsychotic (%)** | 10.3 | 5.1 | 6.3 | 5.7 | 7.9 | - |
| **CPZ Equivalent Dose<sup>^</sup>** | 223.1 (144.2) | 218.0 (210.1) | 238.3 (174.9) | 145.9 (109.7) | 214.5 (175.6) | - |
| **SOPS** |  |  |  |  |  |  |
| Positive | 12.56 (3.9) | 11.74 (4.1) | 12.49 (4.1) | 11.7 (3.6) | 11.53 (3.7) | 1.07* (1.7) |
| Negative | 11.62 (5.7) | 11.78 (5.8) | 12.0 (5.4) | 11.62 (6.6) | 12.05 (6.2) | 1.46* (2.2) |
| Disorganization | 5.46 (3.1) | 5.12 (2.9) | 5.16 (3.1) | 4.56 (3.3) | 5.26 (3.2) | 0.66* (1.2) |
| General | 9.99 (4.3) | 9.49 (4.6) | 9.34 (3.9) | 9.29 (4.5) | 8.82 (4.2) | 1.34* (2.2) |
| **SAS (SD)** | 39.17 (10.7) | 40.51 (11.4) | 37.85 (11.3) | 38.4 (11.0) | 36.94 (10.5) | 22.99* (3.6) |
| **SIAS (SD)** | 28.64 (17.2) | 30.36 (16.2) | 33.41 (16.2) | 32.0 (17.0) | 30.76 (18.0) | 9.04* (8.7) |
| **CDSS (SD)** | 6.35 (5.2) | 6.32 (4.7) | 6.55 (4.8) | 6.57 (5.0) | 5.26 (4.5) | 0.57* (1.4) |

CDSS: Calgary Depression Scale for Schizophrenia; CPZ: Chlorpromazine Equivalent, SAS: Self-Rating Anxiety Scale; SIAS: Social Interaction Anxiety Scale; SOPS: Scale of Psychosis-Risk Symptoms. <sup>+</sup>Healthy control age was significantly different compared to all CHR groups ( $p<.05$ ); <sup>++</sup>Co-Use group was more male than Non-Substance Use, Non-TC, and healthy controls groups ( $p<.05$ ); <sup>+++</sup>Racial distribution differed between healthy control and non-TC groups ( $p<.05$ ); <sup>++++</sup>Non-Substance Use group was more Hispanic than Co-Use group ( $p<.05$ ). <sup>^</sup>If taking antipsychotic medication. \*Healthy control group had lower symptom severity on all scales compared to all CHR groups ( $p<.05$ ).

### Individuals with Co-Use and Single Substance Use Have Comparable Frequency of Use

There were no differences in cannabis use frequency between the Cannabis Use and Co-Use groups (Supplemental Figure 2A, p>.05) nor in tobacco use frequency between the Tobacco Use and Co-Use groups (Supplemental Figure 2B, p>.05). See supplement.

### Psychosis, Depression, and Anxiety Symptoms Do Not Differ Among CHR Substance Use Groups

We investigated if symptom severity differed by substance use group. Symptom severity was assessed across 7 domains: SOPS Positive, Negative, General, and Disorganization; CDSS, SAS, and SIAS. Healthy controls had lower symptoms than all CHR substance use groups (all p<.001, Supplement). There were no differences in psychosis symptoms (Figure 1A-D, p>.05, Supplement), anxiety, social anxiety, or depression (Supplemental Figure 6) among the CHR substance use groups.

**Figure 1.**
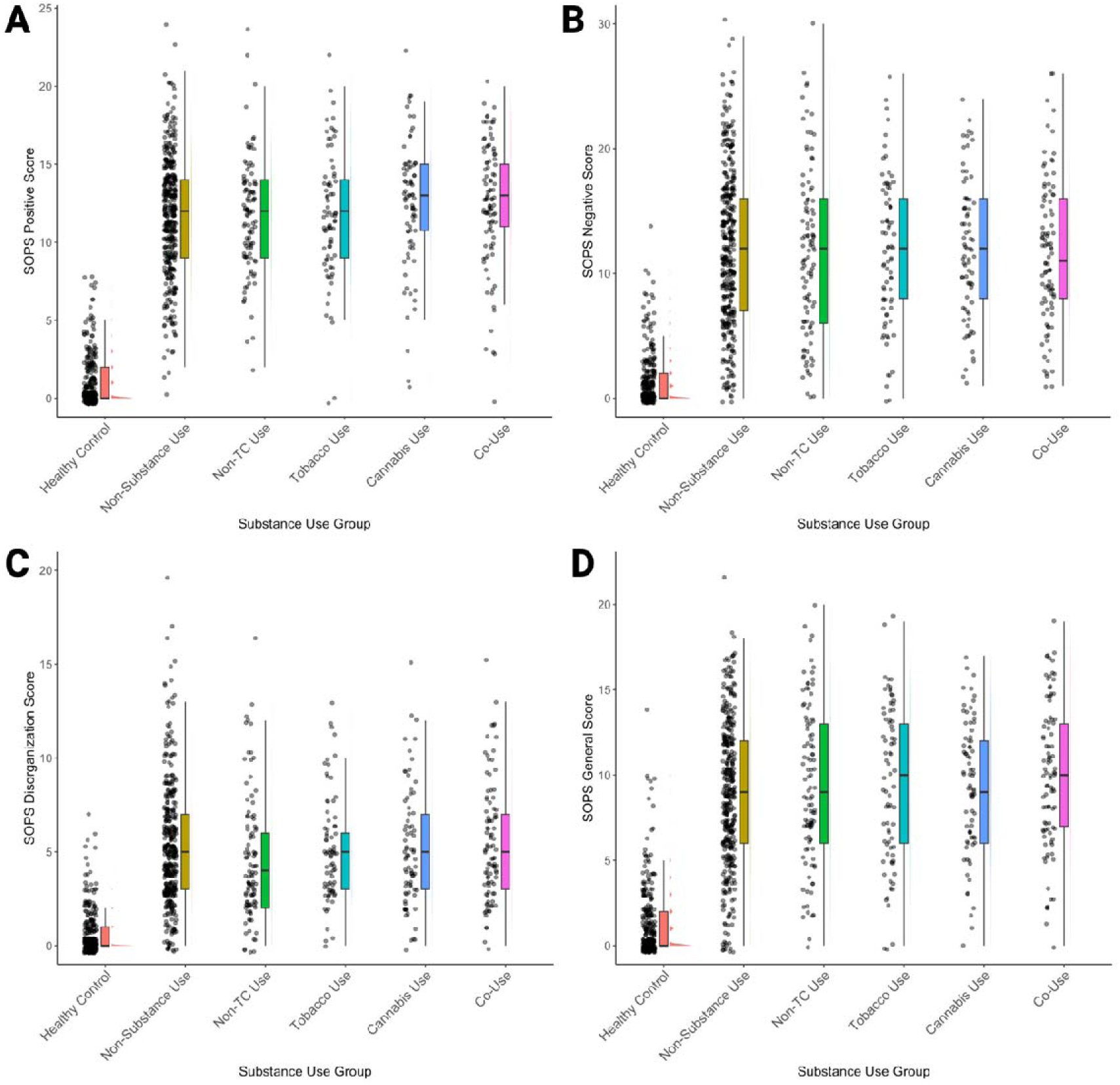
Psychosis Symptoms Do Not Differ Among CHR Substance Use Groups. Shown here are SOPS Positive (1A), Negative (1B), Disorganization (1C), and General (1D) Scores across different substance use groups, including Cannabis Use, Co-Use, Non-Substance Use, Non-TC Use, Tobacco Use, and Healthy Controls. Healthy Controls had significantly lower SOPS scores than all other groups (Non-Substance Use, Non-TC Use, Tobacco Use, Cannabis Use, and Co-Use, Bonferroni-corrected p=.05/7 symptom domains=.007, p<.001), but their significance bars have been omitted for simplicity.

### Greater Tobacco and Cannabis Use are Associated with Greater Psychiatric Symptom Severity

In the combined study population (i.e., CHR and healthy controls), more frequent cannabis and tobacco use was associated with greater psychiatric symptom severity on all domains (Bonferroni-corrected p=.05/7 symptom domains=0.007, all p<.001, Figure 2). See supplement for details and analyses in CHR group alone (Supplemental Figure 7). In linear regression models, diagnosis and sex were significant predictors of psychiatric symptom severity (Supplemental Results).

**Figure 2.**
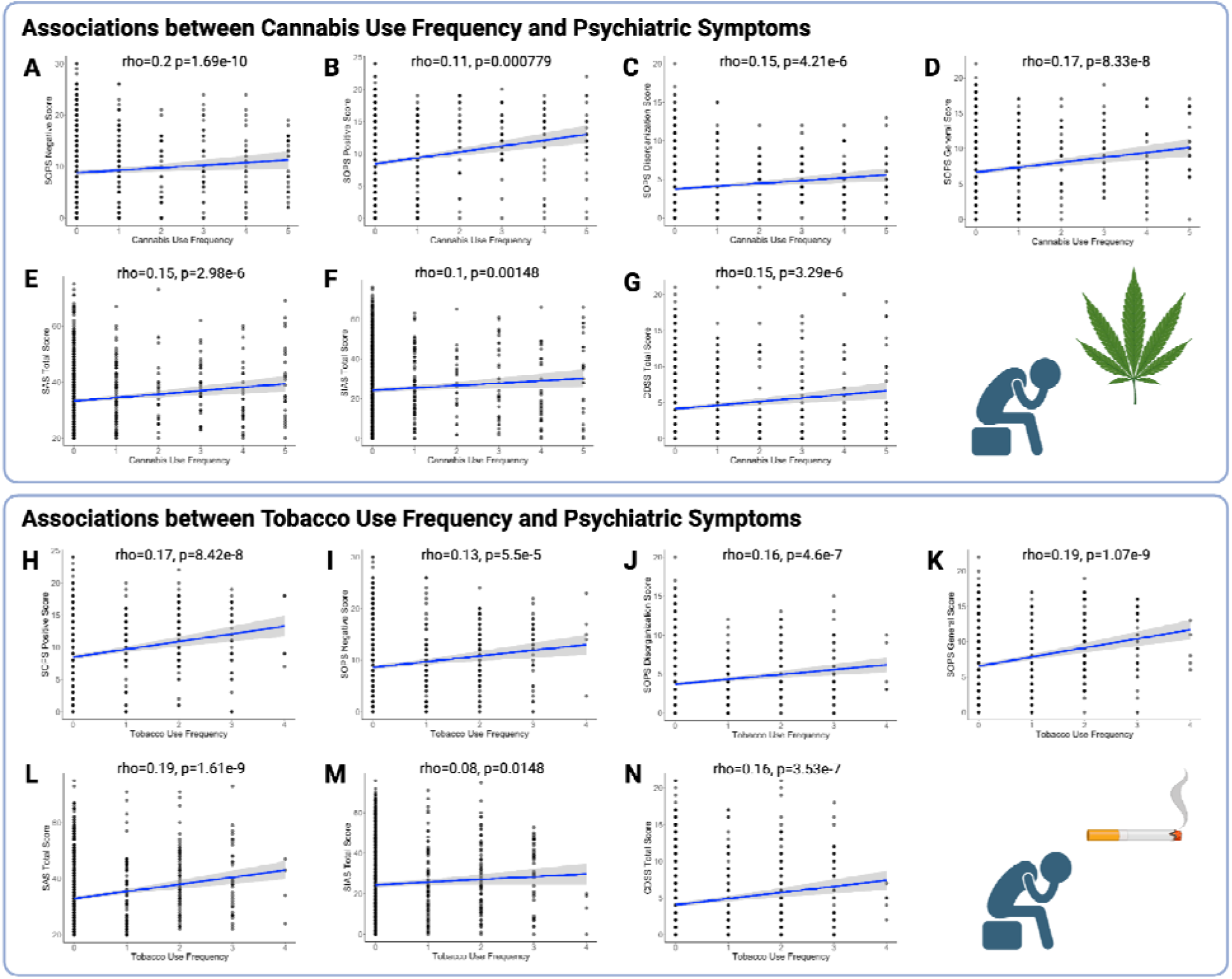
Cannabis and Tobacco Use Frequency Are Associated with Higher Psychiatric Symptom Severity. Across the combined sample (CHR and Healthy Controls), more frequent cannabis use was associated with worsened psychiatric symptoms of psychosis across all SOPS domains (positive, 2A; negative, 2B; disorganization, 2C; general, 2D), anxiety (2E), social anxiety (2F), and depression (2G), Bonferroni-corrected p=.05/7 symptom domains=.007. Similarly, more frequent tobacco use was also associated with worsened psychiatric symptom of psychosis (positive, 2H; negative, 2I; disorganization, 2J; general, 2K) and anxiety (2L), social anxiety (2M), and depression (2N), Bonferroni-corrected p=.05/7 symptom domains=.007. These results suggest a consistent association between more frequent substance use and greater psychiatric symptom burden in this population. Relationships between cannabis and tobacco frequency in the CHR group alone are reported in the Supplement.

### Diagnosis and Tobacco Use Predict Cannabis Use

Linear regression models were used to predict cannabis use frequency based on age, sex, site, diagnosis, tobacco frequency, and diagnosis*tobacco use frequency (F(12,996)=13.23, p<.001). Male sex (Estimate=-0.189, SE=0.070, t=-2.695, p=.0072), CHR diagnosis (Estimate=0.191, SE=0.0836, t=2.287, p=.022), and the Georgia site (Estimate=0.397, SE=0.1434, t=2.768, p=.0057) were significant predictors of cannabis use. The diagnosis*tobacco use interaction (Estimate=0.371. SE=0.146, t=2.537, p=.011) predicted more frequent cannabis use such that CHR participants who used tobacco more frequently also used cannabis more frequently (Supplemental Figure 8).

### Diagnosis and Cannabis Use Predict Tobacco Use

Linear regression models were used to predict tobacco use frequency based on age, sex, site, diagnosis, cannabis use frequency, and diagnosis*cannabis use frequency (F(12,996)=15.97, p<.001). Older age (Estimate=0.0262, SE=0.0058, t=-4.51, p<.001), CHR diagnosis (Estimate=0.224, SE=0.058, t=3.866, p<.001), male sex (Estimate=-0.109, SE=0.0489, t=-2.225, p=.026), and the diagnosis*cannabis use frequency interaction (Estimate=0.157, SE=0.0654, t=2.404, p=.016) were predictors of tobacco use such that CHR participants with more frequent cannabis use also had more frequent tobacco use (Supplemental Figure 8). The Atlanta, New York, North Carolina, and Calgary sites predicted greater tobacco use (see Supplement).

### Survival Analyses

A total of 838 participants (CHR and healthy controls) provided data for survival analyses. Sample details in Supplement. In separate analyses, higher baseline cannabis use (*HR* = 1.27, 95% CI [1.10–1.45], *p* <.001, Supplemental Figure 9) and higher baseline tobacco use (*HR* = 1.29, 95% CI [1.03–1.60], *p* =.02, Supplemental Figure 10) were associated with higher risk of conversion to psychosis. Details in Supplement.

### Tobacco and Cannabis Co-Use is Associated with Higher Risk of Conversion to Psychosis

When we defined tobacco and cannabis use categorically (no use, tobacco use only, cannabis use only, co-use), co-use of cannabis and tobacco was associated with higher risk of conversion compared to no use of either substance (*HR* = 2.53, 95% CI [1.44–4.45], *p* =.001, Figure 3). Cannabis use only (*HR* = 1.54, 95% CI [0.77–3.07], *p* =.22) and tobacco use only (*HR* = 1.69, 95% CI [0.79–3.64], *p* =.18) did not significantly predict conversion compared to no use of either substance. Age had a trending reduced risk of conversion (*HR* = 0.95, 95% CI [0.90– 1.00], *p* =.06), but sex was not associated with conversion.

**Figure 3.**
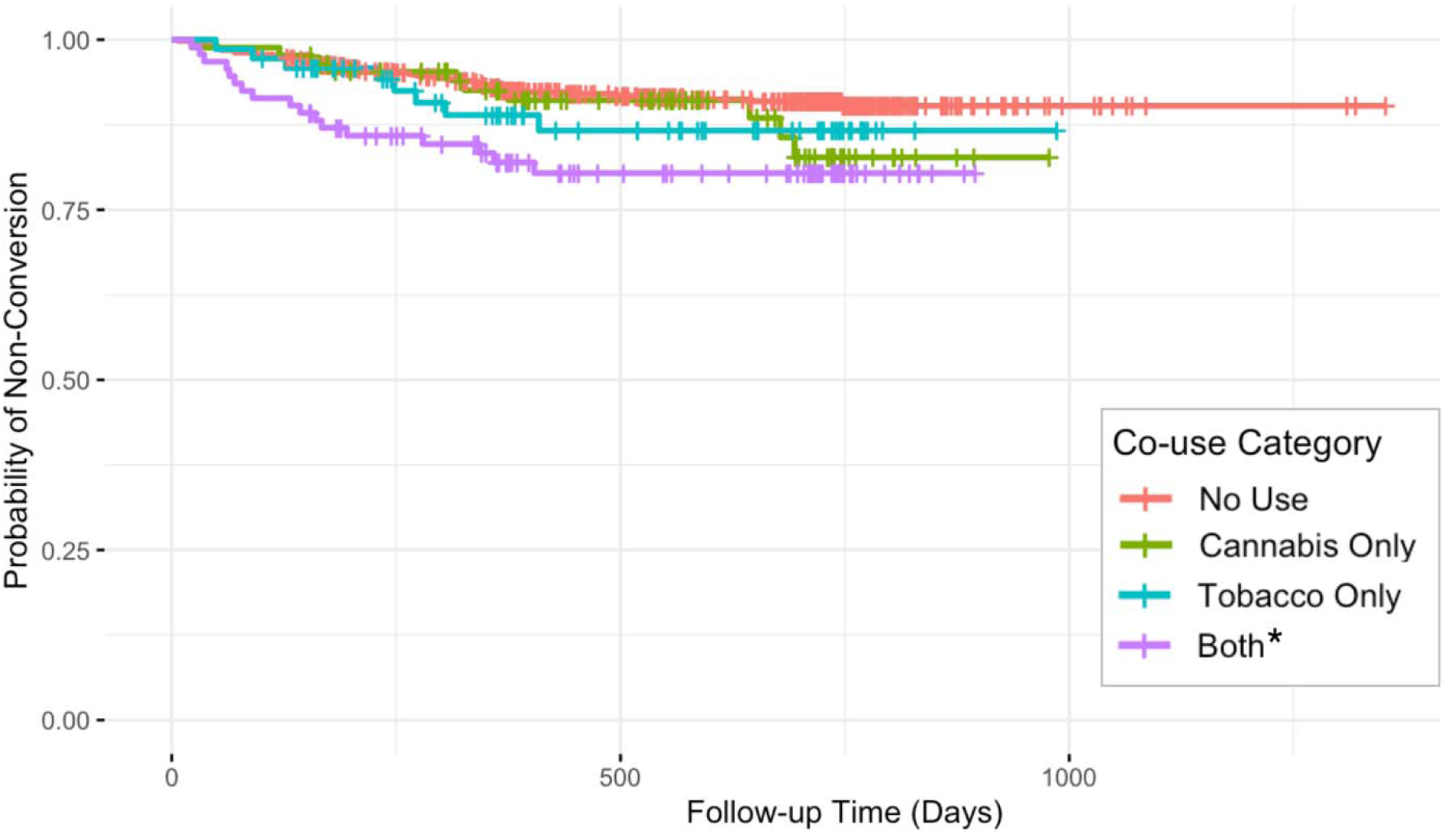
Tobacco and Cannabis Co-Use is Associated with Higher Risk of Conversion to Psychosis. When we defined tobacco and cannabis use categorically (no use, tobacco use only, cannabis use only, co-use), co-use of cannabis and tobacco was associated with higher risk of conversion compared to no use of either substance (*HR* = 2.53, 95% CI [1.44–4.45], *p* =.001). Cannabis use only (*HR* = 1.54, 95% CI [0.77–3.07], *p* =.22) and tobacco use only (*HR* = 1.69, 95% CI [0.79–3.64], *p* =.18) did not significantly predict conversion compared to no use of either substance. Age had a trending reduced risk of conversion (*HR* = 0.95, 95% CI [0.90– 1.00], *p* =.06), but sex was not associated with conversion. Kaplan-Meier survival curves for time to conversion to psychosis by categorial co-use of cannabis and tobacco (None, Cannabis Only, Tobacco Only, Both). The curve is plotted for the purpose of descriptive survival patterns and was not adjusted for age or sex.

### Heavy Co-Use is Associated with the Highest Risk of Conversion to Psychosis

To investigate dose-response relationships of tobacco and cannabis use on conversion to psychosis, we categorized the intensity of cannabis and tobacco use (no use, light use, heavy use for each substance, see Supplemental Methods). Light tobacco use only (HR = 3.08, 95% CI: 1.21–7.85, *p* = 0.018), light cannabis use and heavy tobacco use (HR = 2.86, 95% CI: 1.29– 6.35, *p* = 0.010), and heavy cannabis use and heavy tobacco use (HR = 3.63, 95% CI: 1.53– 8.63, *p* = 0.003, Figure 4) were associated with higher risk of conversion than no use of either substance. Age had a trending reduced risk of conversion (HR = 0.94, 95% CI: 0.89–1.00, *p* = 0.052), but sex was not associated with conversion.

**Figure 4.**
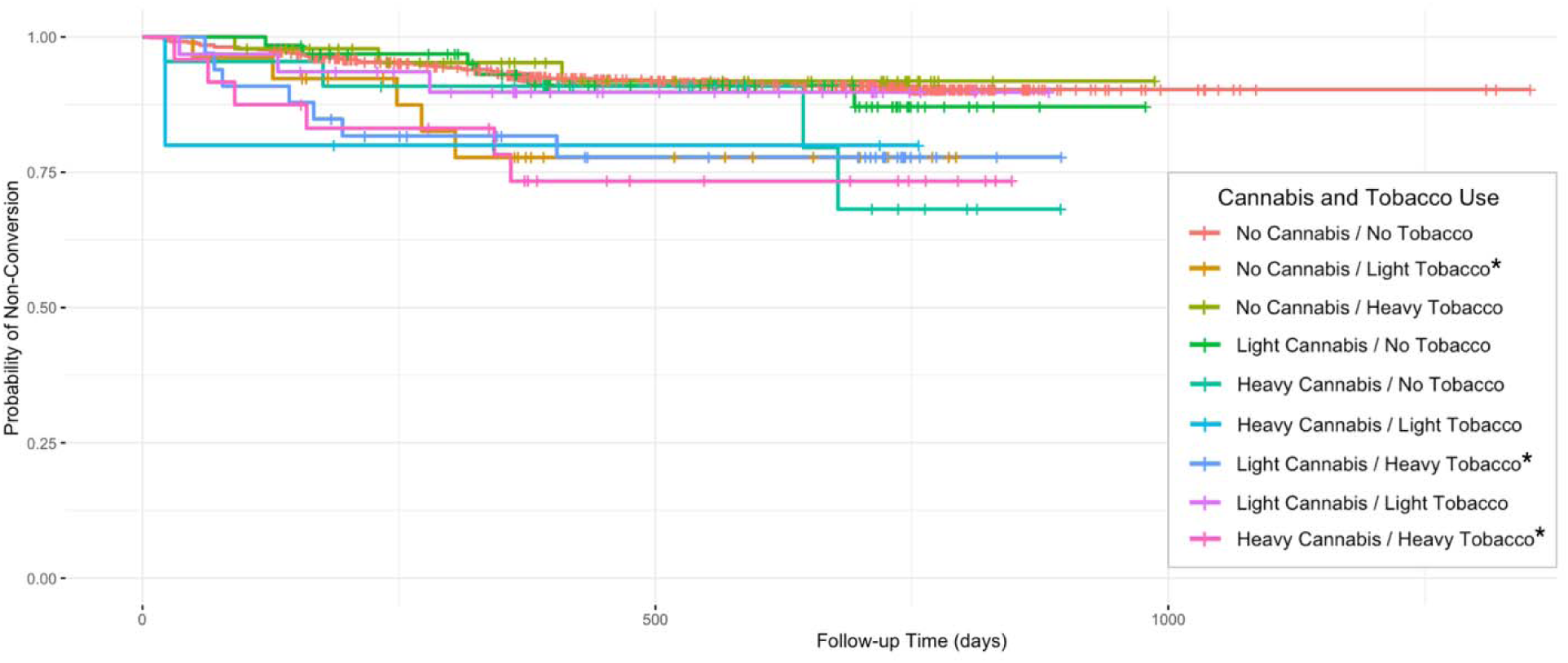
Heavy Co-Use is Associated with the Highest Risk of Conversion to Psychosis. To investigate dose-response relationships of tobacco and cannabis use on conversion to psychosis, we categorized the intensity of cannabis and tobacco use (no use, light use, heavy use for each substance). Light tobacco use only (HR = 3.08, 95% CI: 1.21–7.85, *p* = 0.018), light cannabis use and heavy tobacco use (HR = 2.86, 95% CI: 1.29–6.35, *p* = 0.010), and heavy cannabis use and heavy tobacco use (HR = 3.63, 95% CI: 1.53–8.63, *p* = 0.003) were associated with higher risk of conversion than no use of either substance. Age had a trending reduced risk of conversion (HR = 0.94, 95% CI: 0.89–1.00, *p* = 0.052), but sex was not associated with conversion. Kaplan-Meier survival curves for time to conversion to psychosis by categorial cannabis x tobacco interaction groups (None/Low/High combinations). The curve is plotted for the purpose of descriptive survival patterns and was not adjusted for age or sex.

### Cannabis Use Frequency is Associated with Higher Risk of Conversion when Controlling for Tobacco Use

To isolate the contributions of cannabis and tobacco use individually on conversion to psychosis, we performed analyses using cannabis or tobacco use frequency as a predictor while controlling for use of the other substance. Higher frequency of cannabis use was associated with higher risk of conversion to psychosis (*HR* = 1.22, 95% CI [1.05–1.43], *p* =.009) when controlling for tobacco use. However, tobacco use was no longer significant (*HR* = 1.15, 95% CI [0.90– 1.46], *p* =.26) when controlling for cannabis use. Age had a trending reduced risk of conversion (*HR* = 0.95, 95% CI [0.90–1.01], *p* =.08), but sex was not associated with conversion.

## Discussion

This is the first investigation of tobacco and cannabis co-use in individuals at CHR for psychosis. We observed associations between baseline tobacco and cannabis co-use and psychiatric symptom severity and increased risk of conversion to psychosis. More frequent tobacco and cannabis use was associated with greater symptoms of psychosis, anxiety, and depression. However, psychiatric symptom severity did not differ between substance use groups (i.e., CHR individuals with Co-Use did not have more severe symptoms than with isolated tobacco or cannabis use). These results were contrary to our hypothesis and suggest that tobacco and cannabis use do not have additive effects on clinical symptomatology. However, we also did not observe any beneficial effects of tobacco or cannabis use on symptoms.

Our findings dovetail with mixed reports in the literature. Nicotine transiently improves cognitive deficits and negative symptoms in schizophrenia, likely via agonism of nicotinic acetylcholine receptors to increase dopamine and glutamate transmission in the brain.^7,28,29^ However, heavy nicotine use has been associated with worse outcomes in chronic psychosis, as individuals who smoke more heavily have more severe positive and negative symptoms, more frequent hospitalizations, and higher relapse rates.^7^ Cannabis use exhibits a similarly paradoxical profile: acutely, some individuals report anxiolytic or mood-elevating effects from cannabis, yet sustained use is associated with worsened symptom severity, poorer treatment adherence, and poorer functional outcomes.^11^ Yet, prior research observed that CHR individuals who used cannabis had lower social anhedonia and higher social engagement compared to those who did not use cannabis.^30^

Another critical aspect is why individuals at risk for psychosis are also at greater risk for substance use. Epidemiologically, those who develop schizophrenia are 4–5 times more likely to have had substance use disorders in their youth than the general population.^30^ Similarly, in CHR cohorts, problematic substance use is extremely common (affecting 22% to >50% of CHR individuals) and significantly higher than in age-matched healthy peers. One proposed mechanism is the self-medication hypothesis: adolescents in the prodromal phase often experience subthreshold symptoms of psychosis and may turn to substances to cope with these distressing experiences.^11^ Another important factor is shared neurobiological and genetic vulnerability. There is evidence that neural reward pathways (especially dopaminergic circuits) affect both psychosis and addiction risk.^30^ Individuals with heightened dopaminergic reactivity might be more prone to psychotic experiences *and* more sensitive to the reinforcing effects of substances, making them more likely to use substances. Genetic studies have identified common gene variants that increase the risk for both schizophrenia and substance use disorders (including nicotine dependence), suggesting pleiotropic genetic factors influencing both conditions.^30^ Thus, the high co-occurrence of psychosis-risk status and substance use likely arises from a confluence of factors: attempts to quell emerging symptoms, overlapping biological susceptibilities, and lifestyle or social factors that provide access to substances.

Critically, co-use of tobacco and cannabis was associated with a markedly increased risk of conversion to psychosis. Survival analysis revealed that individuals with co-use had more than double the risk of developing a psychotic disorder relative to CHR with no substance use. In a more granular model differentiating light versus heavy use, individuals with heavy co-use had the highest conversion risk – over a 3-fold increase. Our findings indicate that co-use of tobacco and cannabis is a more potent risk factor for psychosis conversion than use of either substance alone. However, the directionality of this relationship cannot be known. One possible interpretation is a synergistic effect, where tobacco might enhance the impact of cannabis use on the brain’s dopaminergic or endocannabinoid systems, thereby amplifying the biological processes underlying psychosis onset. Both substances increase dopamine in mesolimbic brain regions.^31,32^ Rather than being directly causal, it is also possible that tobacco and cannabis co-use reflects an even higher underlying risk for psychosis among individuals already at CHR for psychosis.

However, cannabis use appears to be the primary culprit, as cannabis use predicted conversion to psychosis even after controlling for tobacco use, which was not true of tobacco when we controlled for cannabis use. These results extend prior research on cannabis use in CHR populations. Cannabis use is linked to increased psychosis risk across multiple epidemiological studies. Prior longitudinal research has shown that heavy cannabis use in adolescence substantially raises the likelihood of later developing schizophrenia-spectrum disorders.^33^ Individuals reporting very frequent cannabis use (50+ times by late teens) have up to a six-fold higher odds of a schizophrenia diagnosis in adulthood compared to individuals without substance use.^33^ Likewise, meta-analyses have concluded that, while cannabis is neither a necessary nor sufficient cause of psychosis, it likely contributes to psychosis risk in predisposed individuals.^33^

### Strengths and Limitations

Our study has several notable strengths. This is the first investigation examining tobacco and cannabis co-use in a CHR cohort. Prior work in psychosis risk largely focused on single substances, whereas our approach captures the real-world scenario where many young people use tobacco and cannabis together. While many cross-sectional studies assess current or lifetime substance use disorders, few studies characterize current substance use below the level of a disorder. Our sample is large (734 CHR individuals, from an initial pool of 1,014 participants including controls) and drawn from eight sites across North America, enhancing generalizability.

While our study sheds new light on tobacco and cannabis co-use in the CHR period, it has several limitations. Our measures of substance use were limited in detail, relying on self-reported current substance use without data on quantity or use history. Further, substance use data lacked granularity to differentiate effects of simultaneous versus asynchronous use of tobacco and cannabis, which may have differing neurobiological effects. Use of non-tobacco forms of nicotine (i.e., patch, lozenge, vaping, pouches) are not assessed in the AUS/DUS. However, as data were collected 2009–2013, vaping is likely uncommon.^34,35^ We could not determine specifics such as cannabis potency (e.g. THC levels or use of high-potency products), which is important as THC potency has increased since 2013.^36,37^ Finally, the observational design precludes conclusions about causality, and we did not include longitudinal assessments in this analysis. Although co-use predicted conversion to psychosis, we cannot be certain that substance use is driving this outcome. It is possible that a shared third factor (e.g. genetic vulnerability, childhood trauma) leads some CHR individuals to both use substances and develop psychosis.

### Future Directions

Given the scarcity of literature on tobacco and cannabis co-use in psychosis risk, our findings expand several avenues for future research. Replication and extension in independent samples will be important to confirm the heightened conversion risk among individuals who co-use tobacco and cannabis. Future studies should investigate longitudinal relationships between patterns of tobacco and cannabis use and psychiatric symptoms in CHR and psychosis spectrum samples. Ongoing prospective studies of CHR individuals should investigate if relationships between co-use and psychiatric symptoms and conversion persist in more recent samples in the context of shifting trends in cannabis legalization and vaping. Future work should include more nuanced measurement of substance use and interrogation of underlying biological mechanisms related to substance use and psychosis risk. From a clinical perspective, our findings suggest that interventions to reduce substance use in CHR populations are warranted. If co-use indeed contributes to conversion risk, then reducing substance use might delay or prevent psychosis onset in some individuals. In conclusion, our study underscores that tobacco and cannabis co-use is an important, yet understudied, factor in the psychosis prodrome.

## Data Availability Statement

Data available upon reasonable request.

## Data Availability

Data available upon reasonable request.

## Acknowledgments

Barbara Cornblatt and Larry Seidman passed away tragically before submission of this manuscript. Their colleagues wish to honor their contributions to the work posthumously.

## Author Contributions

DB: data curation, formal analysis, investigation, visualization, writing – original draft. SB: formal analysis, investigation, visualization, writing – original draft. RR: conceptualization, investigation, writing – reviewing & editing. JA, CB, KC, TC, RC, MK, DM, DO, WS, MT, EW, SW: funding acquisition, investigation, writing – reviewing & editing. BC, LS: funding acquisition, investigation. RO: data curation, investigation, supervision, writing – reviewing & editing. HBW: conceptualization, data curation, formal analysis, funding acquisition, investigation, methodology, project administration, resources, software, supervision, validation, visualization, writing – original draft, and writing – reviewing & editing.

## References

1. Manseau M, Bogenschutz M. Substance Use Disorders and Schizophrenia. FOC. 2016;14(3):333–342. doi:10.1176/appi.focus.20160008

2. Myles H, Myles N, Large M. Cannabis use in first episode psychosis: Meta-analysis of prevalence, and the time course of initiation and continued use. Aust N Z J Psychiatry. 2016;50(3):208–219. doi:10.1177/0004867415599846

3. Koskinen J, Löhönen J, Koponen H, Isohanni M, Miettunen J. Rate of Cannabis Use Disorders in Clinical Samples of Patients With Schizophrenia: A Meta-analysis. Schizophrenia Bulletin. 2010;36(6):1115–1130. doi:10.1093/schbul/sbp031

4. Whitfield-Gabrieli S, Fischer AS, Henricks AM, et al. Understanding marijuana’s effects on functional connectivity of the default mode network in patients with schizophrenia and co-occurring cannabis use disorder: A pilot investigation. Schizophr Res. 2018;194:70–77. doi:10.1016/j.schres.2017.07.029

5. Ward HB, Beermann A, Nawaz U, et al. Evidence for Schizophrenia-Specific Pathophysiology of Nicotine Dependence. Front Psychiatry. 2022;13. doi:10.3389/fpsyt.2022.804055

6. de Leon J, Diaz FJ. A meta-analysis of worldwide studies demonstrates an association between schizophrenia and tobacco smoking behaviors. Schizophrenia Research. 2005;76(2):135–157. doi:10.1016/j.schres.2005.02.010

7. Quigley H, MacCabe JH. The relationship between nicotine and psychosis. Ther Adv Psychopharmacol. 2019;9:2045125319859969. doi:10.1177/2045125319859969

8. Han B, Aung TW, Volkow ND, et al. Tobacco Use, Nicotine Dependence, and Cessation Methods in US Adults With Psychosis. JAMA Network Open. 2023;6(3):e234995. doi:10.1001/jamanetworkopen.2023.4995

9. Winklbaur B, Ebner N, Sachs G, Thau K, Fischer G. Substance abuse in patients with schizophrenia. Dialogues Clin Neurosci. 2006;8(1):37–43.

10. Hasan A, von Keller R, Friemel CM, et al. Cannabis use and psychosis: a review of reviews. Eur Arch Psychiatry Clin Neurosci. 2020;270(4):403–412. doi:10.1007/s00406-019-01068-z

11. Zammit S, Moore THM, Lingford-Hughes A, et al. Effects of cannabis use on outcomes of psychotic disorders: systematic review. Br J Psychiatry. 2008;193(5):357–363. doi:10.1192/bjp.bp.107.046375

12. McClure EA, Rabin RA, Lee DC, Hindocha C. Treatment Implications Associated with Cannabis and Tobacco Co-Use. Curr Addict Rep. 2020;7(4):533–544. doi:10.1007/s40429-020-00334-8

13. French L, Gray C, Leonard G, et al. Early Cannabis Use, Polygenic Risk Score for Schizophrenia and Brain Maturation in Adolescence. JAMA Psychiatry. 2015;72(10):1002–1011. doi:10.1001/jamapsychiatry.2015.1131

14. Bloomfield MAP, Ashok AH, Volkow ND, Howes OD. The effects of Δ9-tetrahydrocannabinol on the dopamine system. Nature. 2016;539(7629):369-377. doi:10.1038/nature20153

15. Hjorthøj C, Compton W, Starzer M, et al. Association between cannabis use disorder and schizophrenia stronger in young males than in females. Psychol Med. 2023;53(15):7322–7328. doi:10.1017/S0033291723000880

16. Shrivastava A, Johnston M, Terpstra K, Bureau Y. Cannabis and psychosis: Neurobiology. Indian J Psychiatry. 2014;56(1):8–16. doi:10.4103/0019-5545.124708

17. Skumlien M, Langley C, Lawn W, et al. The acute and non-acute effects of cannabis on reward processing: A systematic review. Neuroscience & Biobehavioral Reviews. 2021;130:512–528. doi:10.1016/j.neubiorev.2021.09.008

18. Lawn W, Freeman TP, Pope RA, et al. Acute and chronic effects of cannabinoids on effort-related decision-making and reward learning: an evaluation of the cannabis ‘amotivational’ hypotheses. Psychopharmacology (Berl). 2016;233(19):3537–3552. doi:10.1007/s00213-016-4383-x

19. George TP, Verrico CD, Picciotto MR, Roth RH. Nicotinic modulation of mesoprefrontal dopamine neurons: pharmacologic and neuroanatomic characterization. J Pharmacol Exp Ther. 2000;295(1):58–66.

20. Rabin RA, George TP. A review of co-morbid tobacco and cannabis use disorders: possible mechanisms to explain high rates of co-use. Am J Addict. 2015;24(2):105–116. doi:10.1111/ajad.12186

21. Larson MK, Walker EF, Compton MT. Early signs, diagnosis and therapeutics of the prodromal phase of schizophrenia and related psychotic disorders. Expert Rev Neurother. 2010;10(8):1347–1359. doi:10.1586/ern.10.93

22. Powers AR, Addington J, Perkins DO, et al. Duration of the psychosis prodrome. Schizophrenia Research. 2020;216:443–449. doi:10.1016/j.schres.2019.10.051

23. McGlashan T, Walsh B, Woods S. The Psychosis Risk Syndrome: Handbook For Diagnosis and Follow-Up. 1st ed. Oxford University Press; 2010.

24. Drake R, Mueser K, McHugo G. Clinician rating scales: Alcohol use scale (AUS), drug use scale (DUS), and substance abuse treatment scale (SATS). In: Outcome Assessment in Clinical Practice. First. Williams and Wilkins; 1996:113–116.

25. Zung WWK. A Rating Instrument For Anxiety Disorders. Psychosomatics. 1971;12(6):371–379. doi:10.1016/S0033-3182(71)71479-0

26. Mattick RP, Clarke JC. Development and validation of measures of social phobia scrutiny fear and social interaction anxiety. Behav Res Ther. 1998;36(4):455–470. doi:10.1016/s0005-7967(97)10031-6

27. Addington D, Addington J, Maticka-Tyndale E. Assessing depression in schizophrenia: the Calgary Depression Scale. Br J Psychiatry Suppl. 1993;(22):39–44.

28. Meijer JH, Dekker N, Koeter MW, et al. Cannabis and cognitive performance in psychosis: a cross-sectional study in patients with non-affective psychotic illness and their unaffected siblings. Psychol Med. 2012;42(4):705–716. doi:10.1017/S0033291711001656

29. Sacco KA, Termine A, Seyal A, et al. Effects of cigarette smoking on spatial working memory and attentional deficits in schizophrenia: involvement of nicotinic receptor mechanisms. Arch Gen Psychiatry. 2005;62(6):649–659. doi:10.1001/archpsyc.62.6.649

30. Amir CM, Kapler S, Hoftman G, et al. Substance use in youth at genetic and clinical high risk for psychosis. Preprint posted online December 6, 2022. doi:10.1101/2022.12.01.22282991

31. Oleson EB, Cheer JF. A Brain on Cannabinoids: The Role of Dopamine Release in Reward Seeking. Cold Spring Harb Perspect Med. 2012;2(8):a012229. doi:10.1101/cshperspect.a012229

32. Yin R, French ED. A comparison of the effects of nicotine on dopamine and non-dopamine neurons in the rat ventral tegmental area: an *in vitro* electrophysiological study. Brain Research Bulletin. 2000;51(6):507–514. doi:10.1016/S0361-9230(00)00237-9

33. Arseneault L, Cannon M, Ton JW, Ay RMM. Causal association between cannabis and psychosis: examination of the evidence.

34. Smart R, Caulkins JP, Kilmer B, Davenport S, Midgette G. Variation in cannabis potency and prices in a newly legal market: evidence from 30 million cannabis sales in Washington state. Addiction. 2017;112(12):2167–2177. doi:10.1111/add.13886

35. Caulkins JP. Changes in self-reported cannabis use in the United States from 1979 to 2022. Addiction. 2024;119(9):1648–1652. doi:10.1111/add.16519

36. ElSohly MA, Mehmedic Z, Foster S, Gon C, Chandra S, Church JC. Changes in Cannabis Potency over the Last Two Decades (1995-2014) - Analysis of Current Data in the United States. Biol Psychiatry. 2016;79(7):613–619. doi:10.1016/j.biopsych.2016.01.004

37. Chandra S, Radwan MM, Majumdar CG, Church JC, Freeman TP, ElSohly MA. New trends in cannabis potency in USA and Europe during the last decade (2008–2017). Eur Arch Psychiatry Clin Neurosci. 2019;269(1):5–15. doi:10.1007/s00406-019-00983-5

