## Supplemental Materials for "Cannabis and Tobacco Co-Use Predicts Psychosis in Clinical High Risk Cohorts"

**Supplemental Material**

**Supplemental Methods**

**Participants**

*Inclusion and Exclusion Criteria*

Participants underwent clinical assessment at baseline and every 6 months for two years and upon conversion to psychosis (if applicable) from January 2009 to April 2013. Prior to participation, all participants provided written informed consent (or, if under age 18, informed assent with parental consent) in accordance with the institutional review boards of Beth Israel Deaconess Medical Center, Boston, Massachusetts; Emory University, Atlanta, Georgia; University of Calgary, Alberta, Canada; University of California, Los Angeles; University of California, San Diego; The University of North Carolina at Chapel Hill; Yale University, New Haven, Connecticut; and Zucker Hillside Hospital, New York.

CHR individuals were included if they met the Criteria for the Psychosis Risk Syndrome,^1^ based on the Structured Interview for Prodromal Symptoms (SIPS).^2^ If individuals were younger than 19 years, they were included based on criteria for schizotypal personality disorder or Criteria for the Psychosis Risk Syndrome. Individuals could meet any of four prodromal criteria: 1) attenuated positive symptoms (APS), 2) brief intermittent psychotic symptoms (BIPS), 3) genetic risk and deterioration (GRD), or 4) youth and schizotypy criteria (YS). A CHR participant could meet criteria for multiple categories. The Structured Clinical Interview for DSM was used to exclude psychosis and to identify DSM-IV Axis I or cluster A personality disorders. Anyone with a lifetime Axis I psychotic disorder, estimated IQ less than 70 on both measures of IQ, a central nervous system disorder, or DSM-IV substance dependence in the past 6 months was excluded. Other nonpsychotic DSM-IV disorders were not exclusionary (e.g. depression, substance use disorders) unless they clearly caused or better explained prodromal symptoms. CHR individuals were permitted to take antipsychotic medication as long as they had not developed any psychotic symptoms prior to initiating medication. Healthy controls were not permitted to meet any prodromal criteria, have a history of a psychotic or cluster A personality disorder, or have a family history of a psychotic disorder in a first-degree relative.

**Measures**

*Substance Use*: Current substance use was measured using the Alcohol Use Scale/Drug Use Scale (AUS/DUS), which assesses substance use severity (abstinent, use without impairment, abuse, dependence, dependence with institutionalization) and frequency over the past 30 days for the following substances: tobacco, alcohol, marijuana, cocaine, opiates, phencyclidine (PCP), amphetamines, methylenedioxymethamphetamine (MDMA), gamma-hydroxybutarate, huffing, hallucinogens, and other substances.^3^ Substance use frequency was measured on an ordinal scale for tobacco in cigarettes per day (0 = no use, 1 = occasionally, 2 = less than 10 per day, 3 = 11-25 per day, 4 = more than 25 per day) and for all other substances (0 = no use, 1 = once or twice per month, 2 = 3-4 times per month, 3 = 1-2 times per week, 4 = 3-4 times per week, 5 = almost daily). CHR individuals were categorized into one of 5 groups based on their reported substance use in the past 30 days: 1) Tobacco only; 2) Cannabis only; 3) Tobacco and Cannabis Co-Use; 4) Neither Tobacco nor Cannabis (Non-TC users); and 5) No substance use. Healthy controls had minimal substance use and so were treated as one group. A total of 734 CHR individuals had substance use data and were included in the analysis alongside 280 healthy controls.

For the survival analyses, all individuals (healthy and clinical high-risk) were categorized into one of 4 groups based on their reported substance use in the past 30 days: 1) Tobacco only; 2) Cannabis only; 3) Tobacco and Cannabis Co-Use; 4) Neither Tobacco nor Cannabis. Additionally, all individuals were also categorized based on their patterns of use:

Use of neither substance (cannabis = 0 AND tobacco = 0), light tobacco use only (cannabis = 0 AND tobacco = 1), heavy tobacco use only (cannabis = 0 AND tobacco = 2 OR 3 OR 4), light cannabis use only (cannabis = 1 OR 2 OR 3 AND tobacco = 0), heavy cannabis use only (cannabis = 4 OR 5 AND tobacco = 0), heavy cannabis use and light tobacco use (cannabis = 4 OR 5 AND tobacco = 1), light cannabis use and heavy tobacco use (cannabis = 1 OR 2 OR 3 AND tobacco = 2 OR 3 OR 4), light cannabis use and light tobacco use (cannabis = 1 OR 2 OR 3 AND tobacco = 1) and heavy cannabis use and heavy tobacco use (cannabis = 4 OR 5 AND tobacco = 2 OR 3 OR 4) predicted conversion to psychosis compared to no substance use.

*Psychiatric Symptoms*: Severity of positive, negative, disorganized and general symptoms was rated on the Scale of Psychosis-Risk Symptoms (SOPS).^1^ The SOPS is a clinician-administered assessment tool designed to evaluate early signs of psychosis in individuals at clinical high risk (CHR). Developed as part of the Structured Interview for Prodromal Syndromes (SIPS), the SOPS consists of four symptom domains: positive symptoms (e.g., unusual thought content, suspiciousness, perceptual abnormalities), negative symptoms (e.g., social withdrawal, decreased motivation), disorganized symptoms (e.g., trouble with communication and thinking), and general symptoms (e.g., anxiety, sleep disturbances). Anxiety and depressive symptoms were measured through the Self-Rating Anxiety Scale (SAS),^4^ Social Interaction Anxiety scale (SIAS),^5^ and the Calgary Depression Scale for Schizophrenia (CDSS).^6^ The SAS is a self-report questionnaire designed to assess the severity of anxiety symptoms in individuals.^4^ The SIAS is a self-report questionnaire that assesses anxiety experienced in social interactions.^5^ The CDSS is a clinician-administered tool designed to assess depressive symptoms specifically in individuals with schizophrenia.^6^

**Supplemental Results**

*Individuals with Co-Use and Single Substance Use Have Comparable Frequency of Use*

There were no differences in cannabis use frequency between the Cannabis Use and Co-Use groups (H(1)=0.77, p=.38, Supplemental Figure 2A) and no differences in tobacco use frequency between the Tobacco Use and Co-Use groups (H(1)=1.92, p=.17 Supplemental Figure 2B). Alcohol use did not differ between Tobacco Use, Cannabis Use and Co-Use groups (H(2)=0.40, p=.82, Supplemental Figure 3).

*Demographic Differences in Tobacco and Cannabis Use*

Alcohol use frequency did not differ among Tobacco Users, Cannabis Users, and Co-Users (H(2)=0.39899, p=.82 Supplemental Figure 3). Across all subjects, men had a higher frequency of cannabis use (H(1)=8.412, p=.0037) and tobacco use (H(1)=10.17, p=.0014) than women (Supplemental Figures 4-5).

*Healthy Controls have Lower Symptoms than All CHR Substance Use Groups*

We investigated if symptom severity differed by substance use group. Symptom severity was assessed across 7 domains: SOPS Positive, Negative, General, and Disorganization; CDSS, SAS, and SIAS. Healthy controls had lower psychosis symptom severity than all CHR substance use groups (Bonferroni-corrected p=.05/7 symptom domains=0.007, SOPS Positive F(5)=425.11, p<.001; SOPS Negative F(5)=157.15, p<.001; SOPS General F(5)=173.01, p<.001; SOPS Disorganization F(5)=109.44, p<.001). Healthy Controls had lower anxiety (SAS, F(5)=97.55, p<.001), social anxiety (SIAS, F(5)=74.49, p<.001), and depression (CDSS, F(5)=68.717, p<.001, Supplemental Figure 6) scores than all CHR substance use groups.

*Psychiatric Symptoms Do Not Differ Among CHR Substance Use Groups*

Among the CHR substance use groups (Tobacco-only, Cannabis-only, Co-Use, Non-TC, No substance use), there were no differences in psychosis symptoms (Bonferroni-corrected p=.05/7 symptom domains=0.007, SOPS Positive F(5)=425.11, p>.05; SOPS Negative F(5)=157.15, p>.05, SOPS General F(5)=173.01, p>.05, SOPS Disorganization F(5)=109.44, p>.05) or in anxiety (F(5)=97.55, p>.05), social anxiety (F(5)=74.49, p>.05), or depression (F(5)=68.717, p>0.05).

*Greater Tobacco and Cannabis Use are Associated with Greater Psychiatric Symptom Severity*

In the combined study population (i.e., CHR and healthy controls), more frequent cannabis and tobacco use was associated with greater psychiatric symptom severity (Bonferroni-corrected p=.05/7 symptom domains=0.007). More frequent cannabis use was associated with greater severity across all four SOPS symptom domains, including positive (Spearman ρ = 0.2, p = 1.69e-10, Figure 2A), negative (Spearman ρ = 0.11, p<.001, Figure 2B), disorganization (Spearman ρ = 0.15, p<.001, Figure 2C), and general symptoms (Spearman ρ = 0.17, p<.001, Figure 2D), as well as elevated anxiety (Spearman ρ = 0.15, p<.001, Figure 2E), social anxiety (Spearman ρ = 0.1, p=.0015, Figure 2F), and depression (Spearman ρ = 0.15, p<.001, Figure 2G).

Similarly, higher tobacco use frequency was associated with higher symptom severity across these same clinical measures (Bonferroni-corrected p=.05/7 symptom domains=0.007), including positive (Spearman ρ = 0.17, p<.001, Figure 2H), negative (Spearman ρ = 0.13, p<.001, Figure 2I), disorganization (Spearman ρ = 0.16, p<.001, Figure 2J), and general psychosis symptoms (Spearman ρ = 0.19, p<.001, Figure 2K), anxiety (Spearman ρ = 0.19, p<.001, Figure 2L), social anxiety (Spearman ρ = 0.08, p=.015, Figure 2M), and depression severity (Spearman ρ = 3.53e-7, p = 0.16, Figure 2N).

*In Individuals at CHR, Greater Tobacco and Cannabis Use are Associated with Greater Psychiatric Symptom Severity*

In the CHR population alone, more frequent cannabis and tobacco use was associated with greater psychiatric symptom severity (Bonferroni-corrected p=.05/7 symptom domains=0.007). More frequent cannabis use was associated with greater positive symptoms (Spearman ρ = 0.13, p=.0004, Supplemental Figure 7A). However, more frequent cannabis use was not associated with negative (Spearman ρ = 0.015, p=.69, Supplemental Figure 7B), disorganization (Spearman ρ = 0.046, p=.22, Supplemental Figure 7C), or general psychosis symptoms (Spearman ρ = 0.071, p=.056, Supplemental Figure 7D), or anxiety (Spearman ρ = 0.038, p=.32, Supplemental Figure 7E), social anxiety (Spearman ρ = 0.005, p=.89, Supplemental Figure 7F), or depression (Spearman ρ = 0.061, p=.10, Supplemental Figure 7G).

Higher tobacco use frequency was associated with higher anxiety severity (Spearman ρ = 0.106, p=.0054, Supplemental Figure 7L, Bonferroni-corrected p=.05/7 symptom domains=0.007). Tobacco use frequency was not associated with positive (Spearman ρ = 0.06, p=.10, Supplemental Figure 7H), negative (Spearman ρ = 0, p=.99, Supplemental Figure 7I), disorganization (Spearman ρ = 0.047, p=.20, Supplemental Figure 7J), or general psychosis symptoms (Spearman ρ = 0.093, p=.014, Supplemental Figure 7K), or social anxiety (Spearman ρ = 0.043, p=.26, Supplemental Figure 7M), or depression severity (Spearman ρ = .061, p=.10, Supplemental Figure 7N).

To examine relationships with cannabis use frequency in the CHR sample, we ran a linear regression model predicting cannabis use frequency based on age, sex, site, and tobacco use frequency without the effect of diagnosis (F(10,723)=11.52, p<.001). In this model of only individuals at CHR, male sex (Estimate= -0.228, SE=0.091, t=-2.514, p=.012), more frequent tobacco use (Estimate=0.412, SE=0.050, t=8.182, p<.001), and the Georgia site (Estimate=0.5792, SE=0.1886, t=3.071, p=.0022) predicted more frequent cannabis use.

To examine relationships with tobacco use frequency within the CHR sample, we ran a linear regression model predicting tobacco frequency based on age, sex, site, and cannabis use frequency without the effect of diagnosis (F(10,723)=12.31, p<.001). In this model of only individuals at CHR, older age (Estimate=0.0284, SE=0.007939, t=3.582, p<.001), male sex (Estimate= -0.132, SE=0.06425, t=-2.049, p=.041), more frequent cannabis use (Estimate=0.206, SE=0.0251, t=8.182, p<.001), the Georgia (Estimate=0.2843, SE=0.1337, t=2.126, p=.034), New York (Estimate=0.2918, SE=0.1252, t=2.332, p=.020), North Carolina (Estimate=0.3266, SE=0.1252, t=2.608, p=.0093), and Calgary sites (Estimate=0.2935, SE=0.109, t=2.686, p=.0074) were significant predictors of more frequent tobacco use.

*Diagnosis and Sex Predict Psychosis Symptom Severity*

We performed linear regression models predicting psychosis symptom severity based on age, sex, study site, cannabis use frequency, tobacco use frequency, diagnosis (CHR or healthy control), the diagnosis*cannabis frequency interaction, the diagnosis*tobacco frequency interaction, and the cannabis frequency*tobacco frequency interaction.

In a model predicting SOPS positive symptoms (F(15,992)=150.3, p<.001), CHR diagnosis (Estimate=10.84, SE=0.260, t=41.768 p<0.001) and the Georgia site (Estimate=1.6586, SE=0.43105, t=3.848, p<.001) predicted higher scores, while the San Diego site predicted lower scores (Estimate= -1.32557, SE=0.39941, t=-3.319, p<.001).

In a model predicting SOPS negative symptoms (F(15,978)=57.12, p<.001), CHR diagnosis (Estimate=10.74, SE=,0.415, t=25.8782, p<.001), while male sex (Estimate=-1.1056, SE=0.337, t=-3.276, p<.001), and the Boston (Estimate=1.771417, SE=0.7086, t=2.5, p=.013) and New York sites (Estimate=2.004661, SE=0.651242, t=3.078, p=.0021) predicted higher scores.

In a model predicting SOPS general symptoms (F(15,976)=72.31, p<.001), CHR diagnosis (Estimate=7.99, SE=0.286, t=27.936, p<.001), female sex (Estimate=1.077, SE=0.233, t=4.626, p<.001), the New York (Estimate=2.42586, SE=0.44979, t=5.393, p<.001) and Connecticut sites (Estimate=2.83278, SE=0.44079, t=6.427, p<.001), and higher cannabis use frequency (Estimate=0.856, SE=0.291, t=2.942, p=.0033) predicted higher scores, while the interaction between higher cannabis use frequency and CHR diagnosis predicted lower general symptom scores (Estimate= -0.685, SE=0.322, t=-2.129, p=.033).

In a model predicting SOPS disorganization symptoms (F(15,979)=37.83, p<.001), CHR diagnosis predicted higher scores (Estimate = 4.521, SE = 0.219, t=20.643, p<.001), while the San Diego (Estimate= -0.887335, SE=0.341993, t=-2.595, p=.0096) and Calgary sites (Estimate= -0.637002, SE=0.316625, t=-2.012, p=.045) predicted lower disorganization scores.

*Diagnosis, Age, Sex, Tobacco and Cannabis Use Frequency Predict Anxiety and Depression Symptoms*

Linear regression models were also used to predict anxiety (SAS), social anxiety (SIAS), and depression (CDSS) based on age, sex, study site, cannabis use frequency, tobacco use frequency, diagnosis (CHR or healthy control), the diagnosis*cannabis frequency interaction, the diagnosis*tobacco frequency interaction, and the cannabis frequency*tobacco frequency interaction.

In a model predicting anxiety severity (F(15,930)=42.35, p<0.001, CHR diagnosis (Estimate=14.632, SE=0.739, t=19.794, p<.001), female sex (Estimate=4.434, SE=0.596, t=7.445, p<.001), the Calgary site (Estimate=4.510211, SE=1.087989, t=4.145, p<.001), and higher cannabis use frequency (Estimate=1.722, SE=0.730, t=2.359, p=.019) predicted greater anxiety.

In a model predicting social anxiety severity, (F(15,924)=27.06, p<0.001, CHR diagnosis (Estimate=23.290, SE=1.272, t=18.311, p<.001) and older age (Estimate=0.473, SE=0.123, t=3.862, p<.001) predicted higher scores.

In a model predicting depression severity, (F(15,969)=27.87, p<0.001, CHR diagnosis (Estimate=5.438; SE=0.321; t=19.951; p<.001), female sex (Estimate=0.654, SE=0.260, t=2.513, p=.012), older age (Estimate=0.163, SE=0.03127, t=5.214, p<.001), and the New York site (Estimate=1.89586, SE=0.50541, t=3.751, p<.001) predicted higher scores, while the North Carolina site (Estimate= -1.06914, SE=0.51524, t=-2.075, p=.038) predicted lower depression scores.

*Diagnosis and Cannabis Use Predict Tobacco Use*

Linear regression models were used to predict tobacco use frequency based on age, sex, site, diagnosis, cannabis use frequency, and diagnosis*cannabis use frequency (F(12,996)=15.97, p<.001). The Georgia (Estimate=0.2324, SE=0.0999, t=2.325, p=.020), New York (Estimate=0.203, SE=0.0933, t=2.177, p=.030), North Carolina (Estimate=0.224073, SE=0.095, t=2.358, p=.0186), and Calgary sites (Estimate=0.2365, SE=0.0853, t=2.771, p=.0057) were all predictors of greater tobacco use frequency.

*Tobacco and Cannabis Use are Associated with Prodromal Diagnostic Criteria*

As an exploratory analysis, we tested if prodromal diagnostic criteria (APS, BIPS, GRD, YS) predicted cannabis use in the CHR group (F(4,730)=3.127, p=.014). Individuals who met GRD criteria had higher cannabis use frequency (Estimate=0.3847, SE=0.1660, t=2.318, p=.021). We then included the prodromal diagnostic criteria into a model alongside age, sex, site, and tobacco use frequency (F(14,719)=8.937, p<.001). In this model, meeting GRD criteria (Estimate=0.346, SE=0.158, t=2.187, p=.029), the Georgia site (Estimate=0.6536, SE=0.1905, t=3.431, p=.00064), and higher tobacco frequency (Estimate=0.4013, SE=0.0505, t=7.939, p<.001) were significant predictors of cannabis use.

We then tested if prodromal diagnostic criteria predicted tobacco use frequency in the CHR sample (F(4,730)=3.09, p=.015). Individuals who met APS (Estimate=-0.3511, SE=0.145, t=-2.415, p=.016) criteria used tobacco more frequently. When we included prodromal diagnostic criteria alongside age, sex, site, and cannabis use frequency (F(14,719)=9.331, p<.001), older age (Estimate=0.0282, SE=0.008235, t=3.423, p<.001), individuals who met APS criteria (Estimate=-0.317, SE=0.2359, t=-2.318, p=.021), and higher cannabis use frequency (Estimate=0.200834, SE=0.0253, t=7.939, p<.001) were significant predictors of tobacco use such that CHR participants who used cannabis more frequently also used tobacco more frequently. In addition, the Georgia (Estimate=0.28805, SE=0.135418, t=2.127, p=.034), New York (Estimate=0.298937, SE=0.125253, t=2.387, p=.017), North Carolina (Estimate=0.339443, SE=0.125407, t=2.707, p=.0070), Calgary (Estimate=0.301906, SE=0.110031, t=2.744, p=.0062), and Connecticut sites (Estimate=0.245233, SE=0.122051, t=2.009, p=.045) were all predictors of greater tobacco use frequency.

***Survival Analyses***

Survival analyses were performed in a subset of 838 participants, including both healthy controls and CHR individuals, with complete data. This subset had a mean age of 18.79 years (SD = 4.43). The majority (55.97%) were male, and 83 of these individuals converted to psychosis at follow up (9.90%). In this sample, 587 individuals reported no tobacco or cannabis use, 72 individuals reported tobacco use only, 86 individuals reported cannabis use only, and 93 individuals reported using both tobacco and cannabis.

*Frequency of Cannabis Use Alone is Associated with Higher Risk of Conversion in Entire Sample*

Higher frequency of cannabis use at baseline was associated with higher risk of conversion to psychosis (HR = 1.27, 95% CI [1.10–1.45], p < .001, Supplemental Figure 9)*.* Age had a trending reduced risk of conversion (HR = 0.95, 95% CI [0.90–1.00], p = .07), and sex was not associated with significant risk of conversion in this model.

*Frequency of Tobacco Use Alone is Associated with Higher Risk of Conversion in Entire Sample*

Higher frequency of tobacco use at baseline was associated with higher risk of conversion to psychosis (HR = 1.29, 95% CI [1.03–1.60], p = .02, Supplemental Figure 10)*.* Age had a trending reduced risk of conversion (HR = 0.95, 95% CI [0.90–1.01], p = .08), and sex was not associated with significant risk of conversion in this model.

**Supplemental Tables & Figures**

**Supplemental Table 1. NAPLS2 Demographics**

|  | | **CHR** | | | **Healthy Control (n=280)** |
| --- | --- | --- | --- | --- | --- |
|  |  | **CHR (n=764)** | **CHR-NC (n=678)** | **CHR-C (n=86)** |  |
| Age, y (SD) | | 18.6 (4.3)* | 18.6 (4.3) | 18.0 (3.6) | 19.7 (4.7) |
| Sex, Male (%) | | 436 (57.1) | 382 (56.3) | 54 (62.8) | 141 (50.4) |
| Race | |  |  |  |  |
|  | First Nations (%) | 13 (1.7) | 12 (1.8) | 1 (1.2) | 4 (1.4) |
|  | East Asian (%) | 19 (2.5) | 18 (2.7) | 1 (1.2) | 15 (5.4) |
|  | Southeast Asian (%) | 15 (2.0) | 12 (1.8) | 3 (3.5) | 7 (2.5) |
|  | South Asian (%) | 20 (2.6) | 17 (2.5) | 3 (3.5) | 8 (2.9) |
|  | Black (%) | 118 (15.4) | 107 (15.8) | 11 (12.8) | 49 (17.5) |
|  | Central/South American (%) | 34 (4.5) | 30 (4.4) | 4 (4.7) | 13 (4.6) |
|  | West/Central Asia and Middle East (%) | 7 (0.9) | 6 (0.9) | 1 (1.2) | 2 (0.7) |
|  | White (%) | 437 (57.2) | 389 (57.5) | 48 (55.8) | 152 (54.3) |
|  | Native Hawaiian or Pacific Islander (%) | 3 (0.4) | 2 (0.3) | 1 (1.2) | 1 (0.4) |
|  | Interracial (%) | 97 (12.7) | 84 (12.4) | 13 (15.1) | 29 (10.4) |
| Hispanic (%) | | 142 (18.6) | 126 (18.6) | 16 (18.6) | 50 (17.9) |
| Taking Antipsychotic (%) | | 57 (7.5) | 48 (7.1) | 9 (10.5) | - |

CHR: clinical high risk; CHR-NC: clinical high risk – nonconverter; CHR-C: clinical high risk – converter; HC: healthy control. Comparisons were made between 1) CHR and healthy controls and 2) between CHR-C and CHR-NC. There were no significant differences in demographic variables between CHR-C and CHR-NC. ***p<.001**

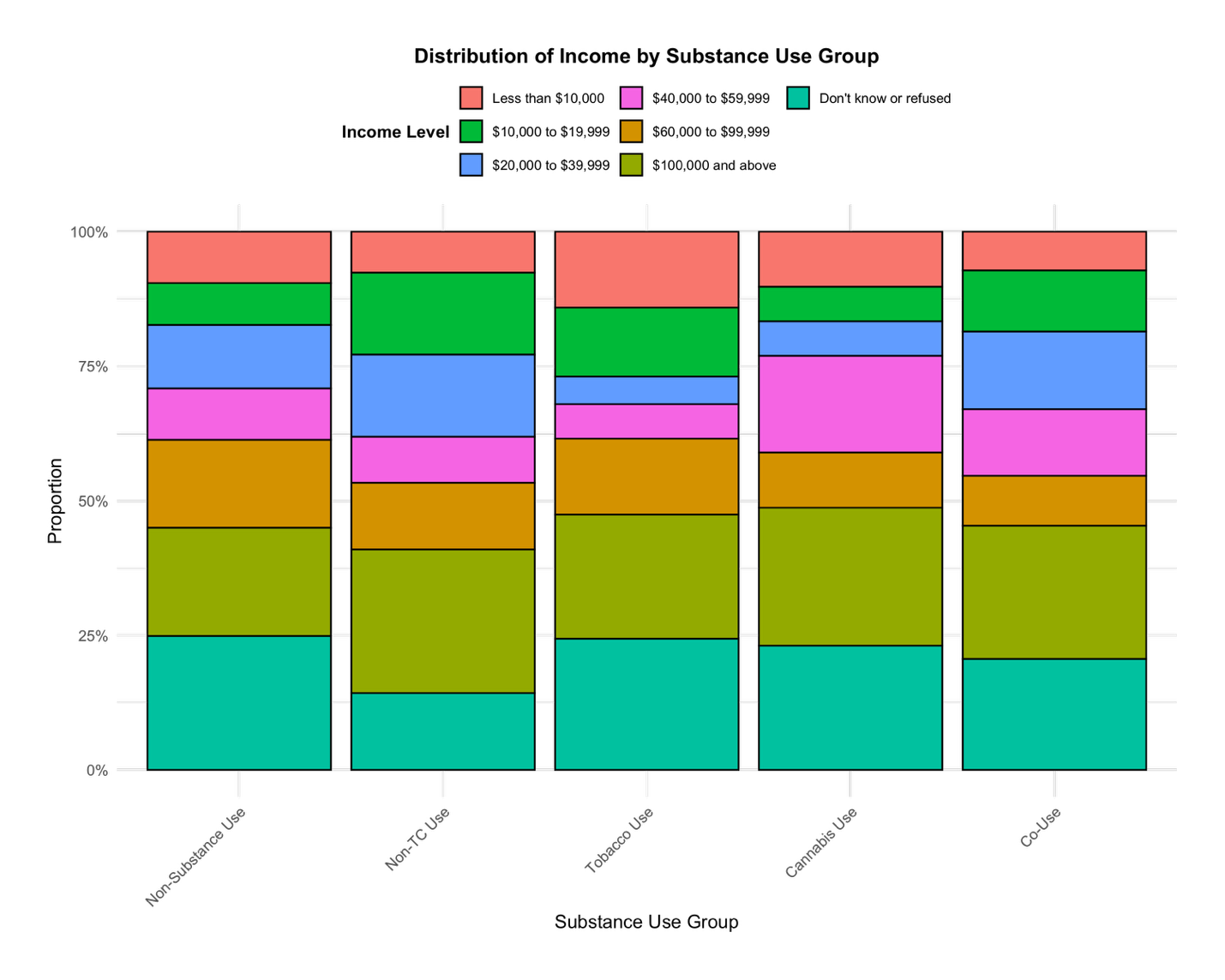

**Supplemental Figure 1. Socioeconomic Status Does Not Differ Across CHR Substance Use Groups.** Socioeconomic status was measured using household income across CHR substance use groups. There were no significant differences in socioeconomic status between groups (p=.48).

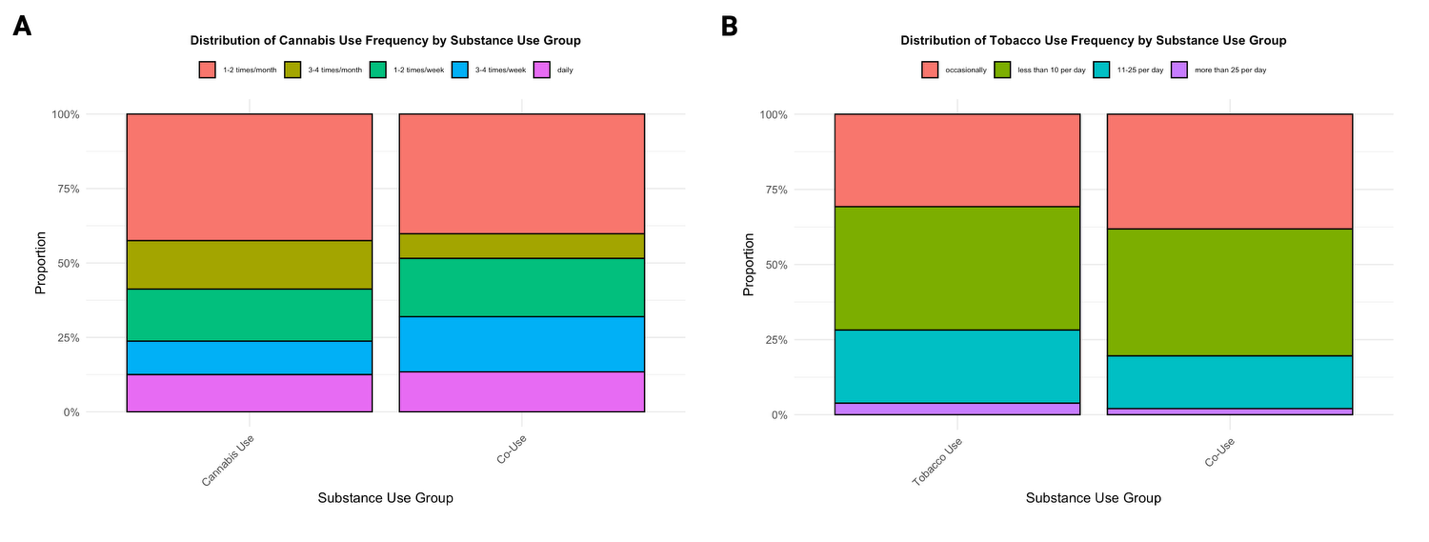

**Supplemental Figure 2. Cannabis and Tobacco Use Frequency Does Not Differ Between Cannabis- or Tobacco Users and Co-Users*.*** Cannabis use frequency as measured by the AUS/DUS did not differ between Cannabis Users and Co-Users (p=.38, 2A). Tobacco use frequency, also measured by the AUS/DUS, did not differ between Tobacco Users and Co-Users (p=.38, 2B).

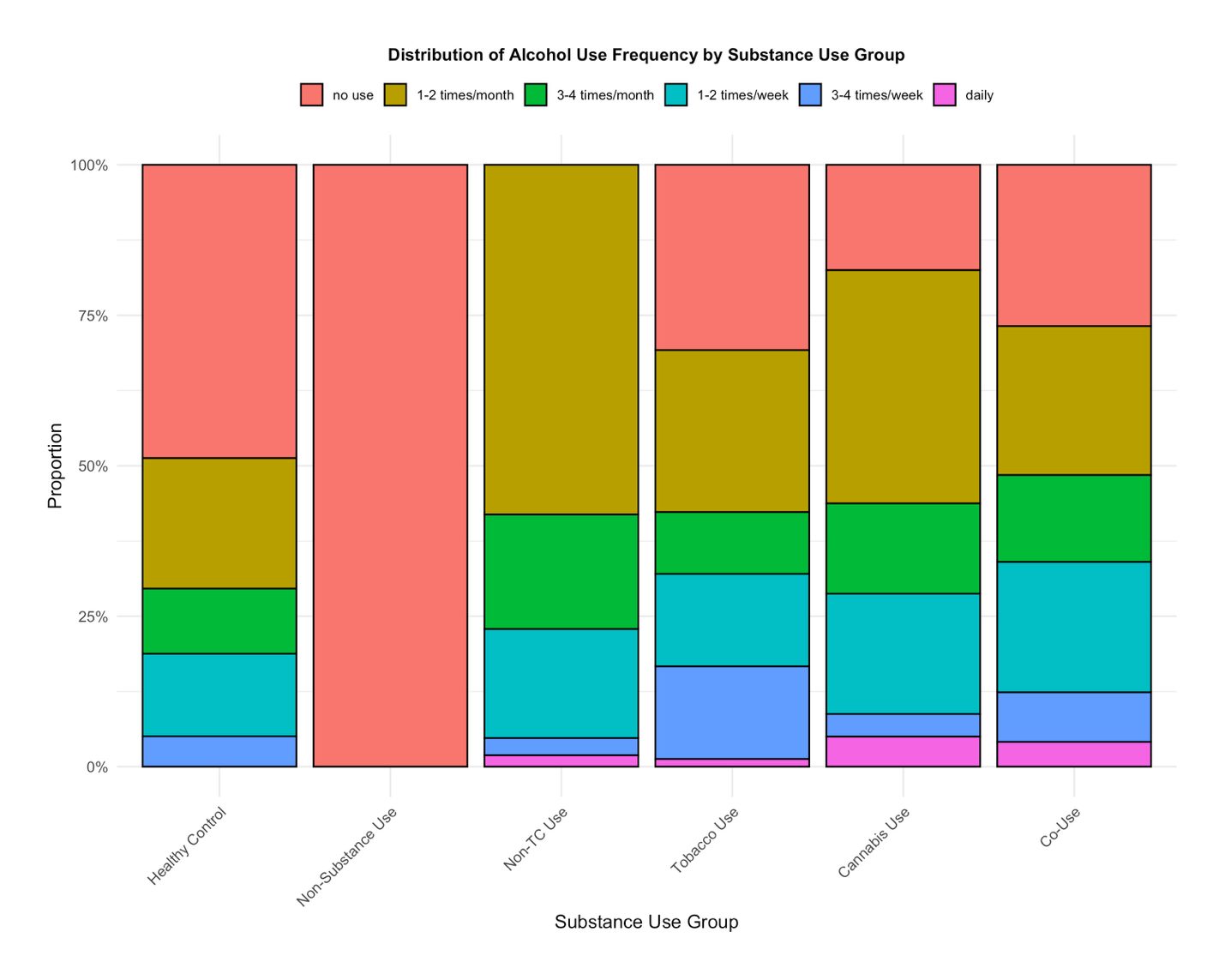
**Supplemental Figure 3. Alcohol Use Frequency Does Not Differ Among Tobacco Users, Cannabis Users, and Co-Users in the Sample.** Alcohol use frequency did not differ among Tobacco Users, Cannabis Users, and Co-Users in the CHR group (p=.82).

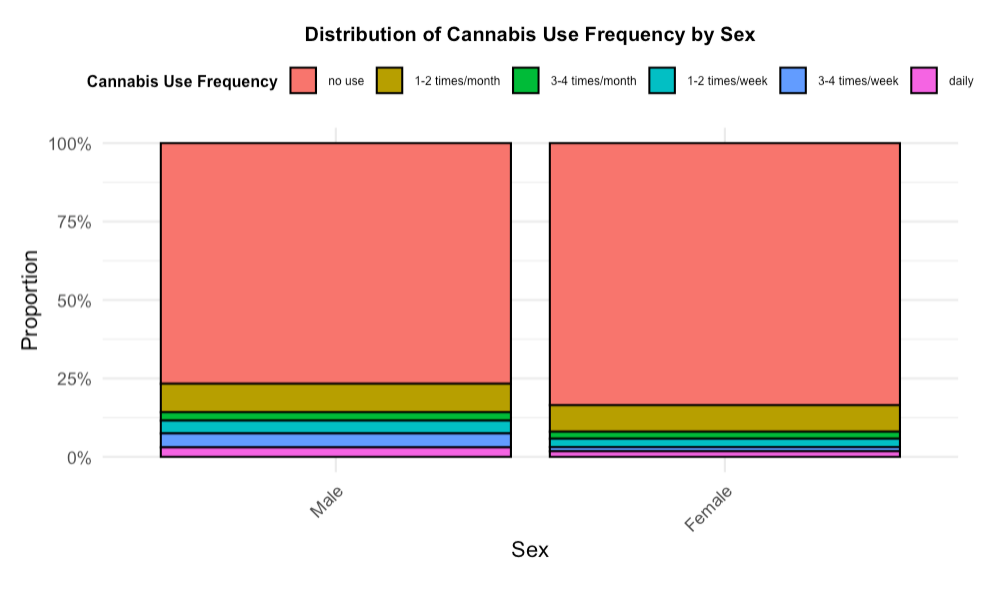

**Supplemental Figure 4. Males Have Higher Cannabis Use Frequency Than Females.** Across the combined sample (CHR and Healthy Controls), males used cannabis more frequently than females (χ²=8.412, df=1, p=.0037), as measured by the AUS/DUS.

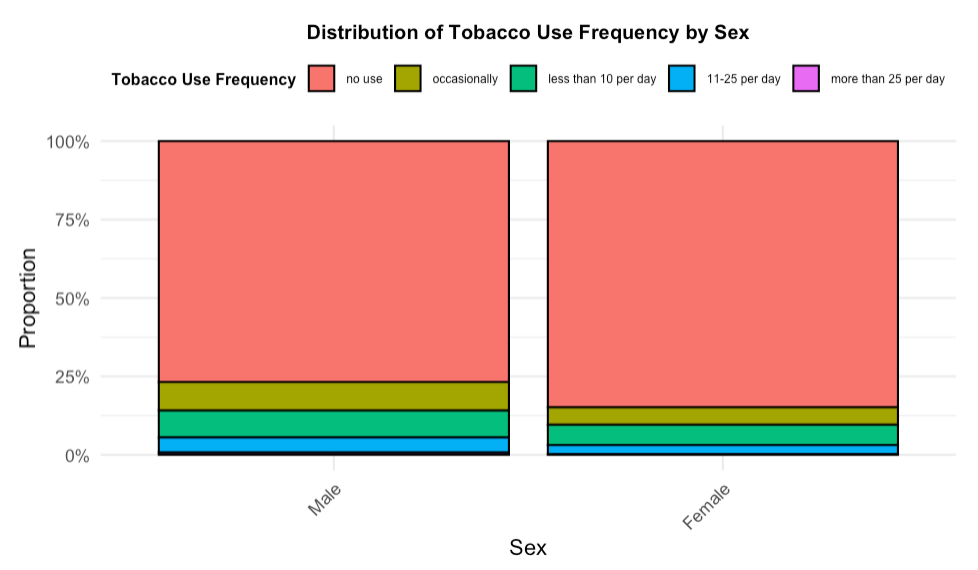
**Supplemental Figure 5. Males Have Higher Tobacco Use Frequency Than Females.** Across the combined sample (CHR and Healthy Controls), males used tobacco more frequently than females (χ²=10.17, df=1, p=.0014), as measured by the AUS/DUS.

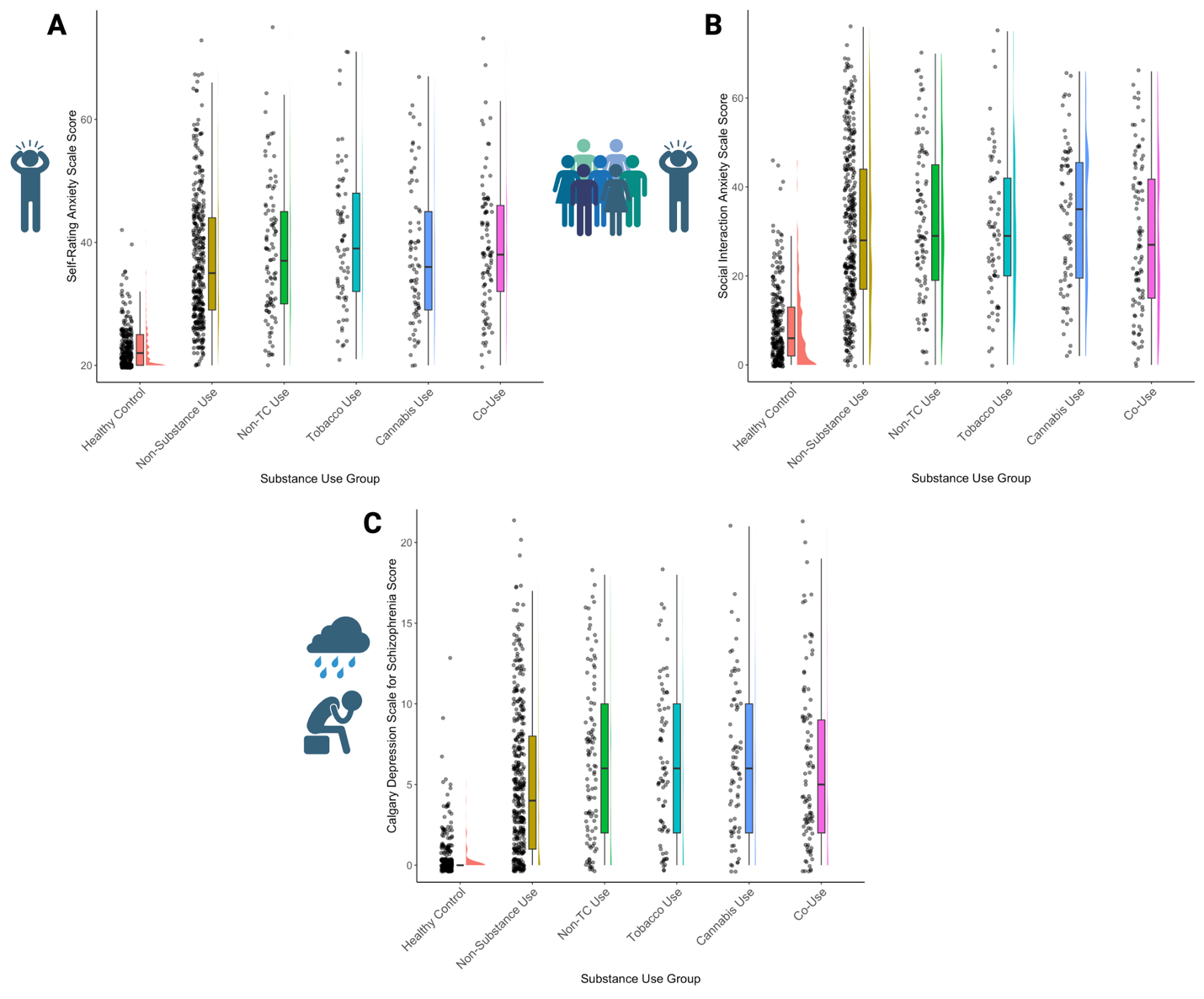

**Supplemental Figure 6. Anxiety and Depression Severity Do Not Differ Across CHR Substance Use Groups.** Anxiety (SAS, 6A), social anxiety (SIAS, 6B), and depression (CDSS, 6C) scores did not differ across CHR substance use groups. Healthy Controls had significantly lower anxiety, social anxiety, and depression scores than all other groups (Non-Substance Use, Non-TC Use, Tobacco Use, Cannabis Use, and Co-Use, Bonferroni-corrected p=.05/7 symptom domains=.007, p<.001), but their significance bars have been omitted for simplicity.

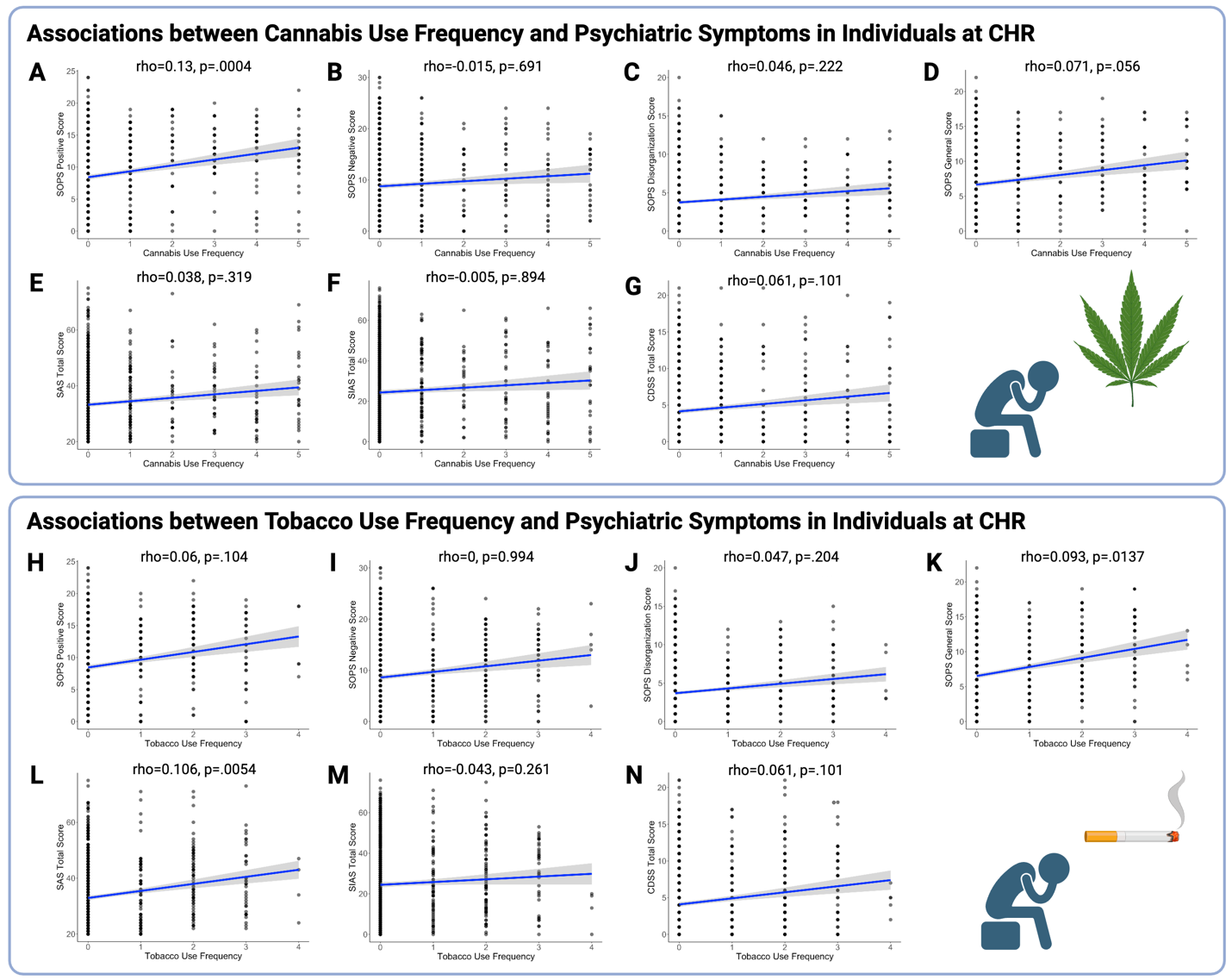

**Supplemental Figure 7. In the CHR Group, More Frequent Cannabis Use is Associated with Higher Positive Symptom Severity, while More Frequent Tobacco use is Associated with Higher Anxiety**. In the CHR population alone, more frequent cannabis use was associated with greater positive symptoms (Spearman ρ = 0.13, p=.0004, Supplemental Figure 7A, Bonferroni-corrected p=.05/7 symptom domains=0.007). However, more frequent cannabis use was not associated with negative (Supplemental Figure 7B), disorganization (Supplemental Figure 7C), or general psychosis symptoms (Supplemental Figure 7D), or anxiety (Supplemental Figure 7E), social anxiety (Supplemental Figure 7F), or depression (Supplemental Figure 7G). More frequent tobacco use was associated with higher anxiety severity (Spearman ρ = 0.106, p=.0054, Supplemental Figure 7L). Tobacco use frequency was not associated with positive (Supplemental Figure 7H), negative (Supplemental Figure 7I), disorganization (Supplemental Figure 7J), or general psychosis symptoms (Supplemental Figure 7K), or symptoms of social anxiety (Supplemental Figure 7M), or depression (Supplemental Figure 7N).

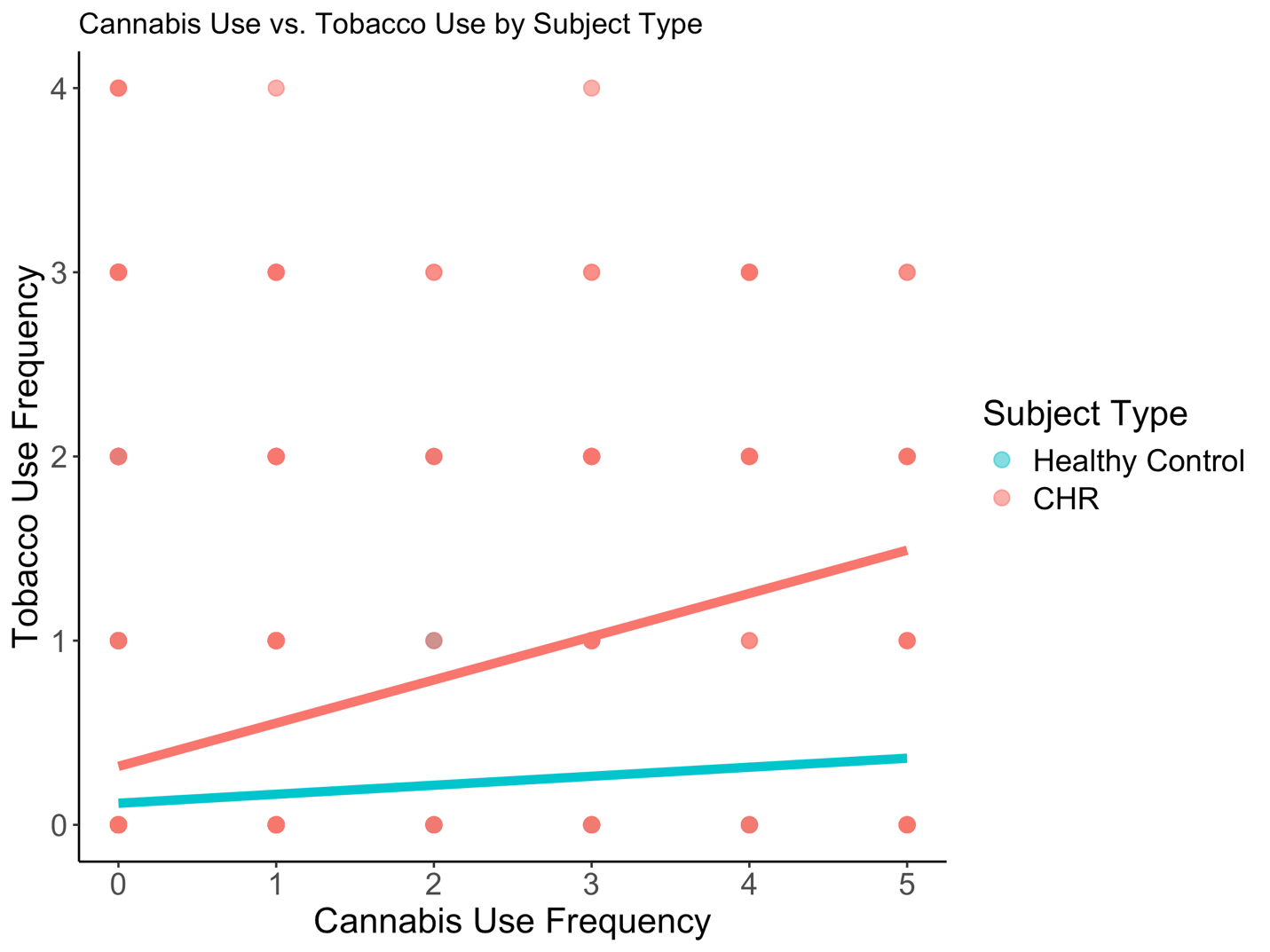

**Supplemental Figure 8. More Frequent Cannabis Use is Associated with More Frequent Tobacco Use.** In a linear regression model to predict tobacco use frequency controlling for age, sex, site, diagnosis, cannabis frequency, and the diagnosis*cannabis use frequency interaction (F(12,996)=15.97, p<.001), older age (Estimate=0.0262, SE=0.0058, t=-4.51, p<0.001), CHR diagnosis (Estimate=0.224, SE=0.058, t=3.866, p<.001), male sex (Estimate=-0.109, SE=0.0489, t=-2.225, p=0.026), and the diagnosis*cannabis use frequency interaction (Estimate=0.157, SE=0.0654, t=2.404, p=.016) were predictors of tobacco use such that CHR participants with more frequent cannabis use also had more frequent tobacco use. The Atlanta, New York, North Carolina, and Calgary sites predicted greater tobacco use (p<.05).

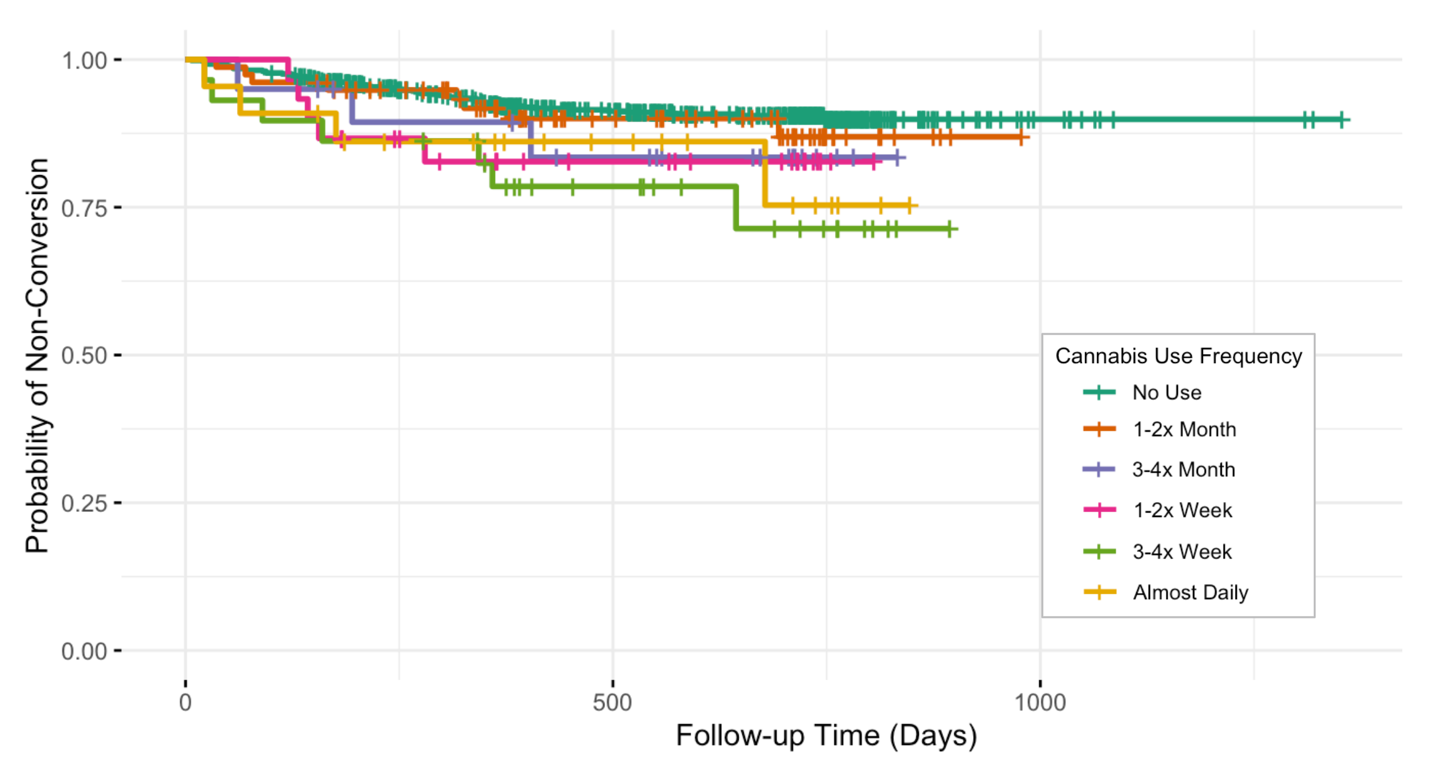

**Supplemental Figure 9. Cannabis Use Frequency is Associated with Higher Conversion Risk.** Higher frequency of cannabis use at baseline was associated with higher risk of conversion to psychosis (HR = 1.27, 95% CI [1.10–1.45], p < .001*).* Age had a trending reduced risk of conversion (HR = 0.95, 95% CI [0.90–1.00], p = .07), but sex was not associated with conversion.

Kaplan-Meier survival curves for time to conversion to psychosis by baseline ordinal cannabis use. The curve is plotted the purpose of descriptive survival patterns and was not adjusted for age or sex.

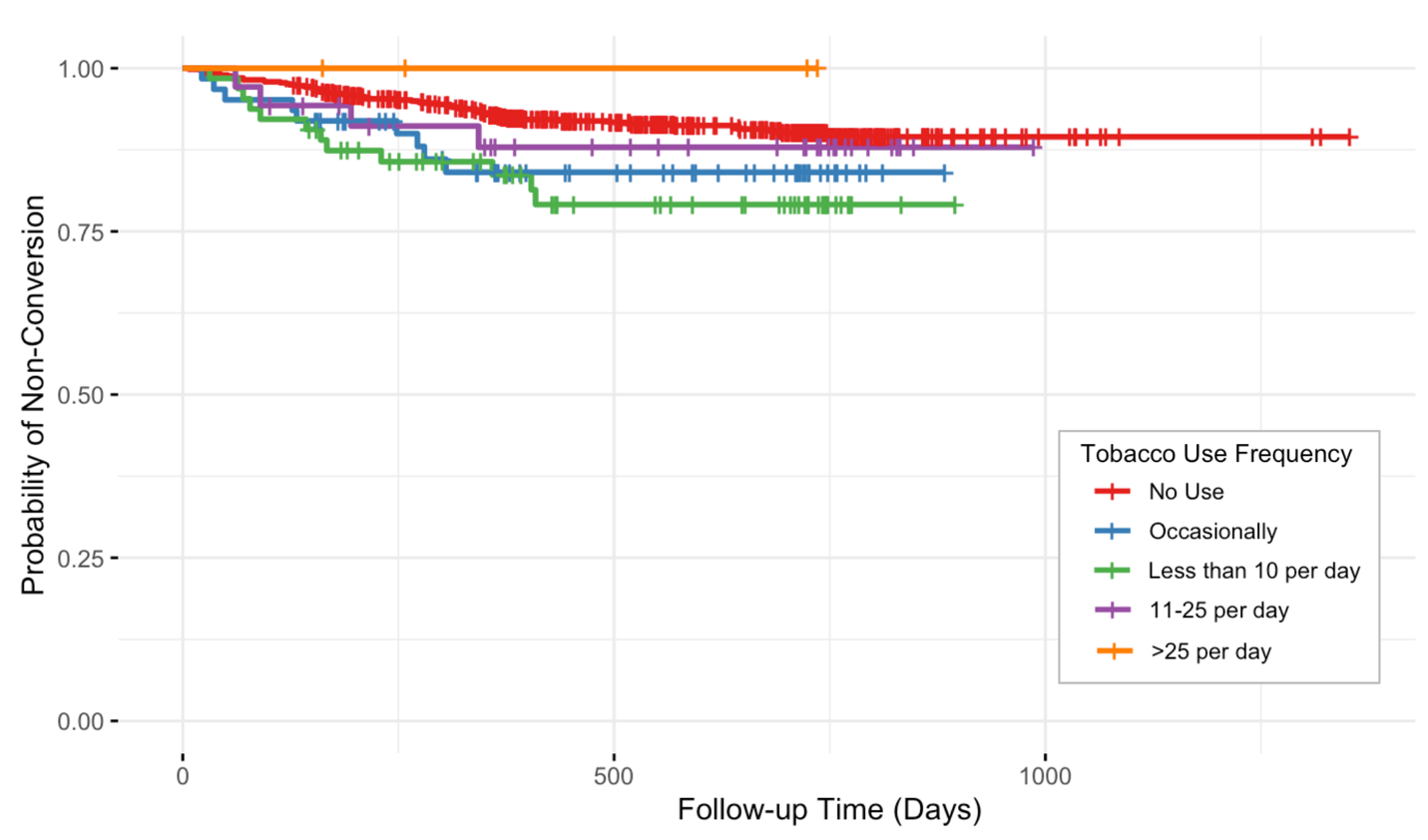

**Supplemental Figure 10. Tobacco Use Frequency is Associated with Higher Conversion Risk.** Higher frequency of tobacco use was associated with higher risk of conversion to psychosis (HR = 1.29, 95% CI [1.03–1.60], p = .02)*.* Age had a trending reduced risk of conversion (HR = 0.95, 95% CI [0.90–1.01], p = .08), but sex was not associated with conversion. Kaplan-Meier survival curves for time to conversion to psychosis by baseline ordinal tobacco use. The curve is plotted for the purpose of descriptive survival patterns and was not adjusted for age or sex.

**References**

1.	McGlashan T, Walsh B, Woods S. *The Psychosis Risk Syndrome: Handbook For Diagnosis and Follow-Up*. 1st ed. Oxford University Press; 2010.

2.	Miller TJ, McGlashan TH, Rosen JL, et al. Prodromal assessment with the structured interview for prodromal syndromes and the scale of prodromal symptoms: predictive validity, interrater reliability, and training to reliability. *Schizophr Bull*. 2003;29(4):703-715. doi:10.1093/oxfordjournals.schbul.a007040

3.	Drake R, Mueser K, McHugo G. Clinician rating scales: Alcohol use scale (AUS), drug use scale (DUS), and substance abuse treatment scale (SATS). In: *Outcome Assessment in Clinical Practice*. First. Williams and Wilkins; 1996:113-116.

4.	Zung WWK. A Rating Instrument For Anxiety Disorders. *Psychosomatics*. 1971;12(6):371-379. doi:10.1016/S0033-3182(71)71479-0

5.	Mattick RP, Clarke JC. Development and validation of measures of social phobia scrutiny fear and social interaction anxiety. *Behav Res Ther*. 1998;36(4):455-470. doi:10.1016/s0005-7967(97)10031-6

6.	Addington D, Addington J, Maticka-Tyndale E. Assessing depression in schizophrenia: the Calgary Depression Scale. *Br J Psychiatry Suppl*. 1993;(22):39-44.
